# Evaluation of the effectiveness of mass drug administration strategies for reducing scabies burden in Monrovia, Liberia: An agent-based modelling approach

**DOI:** 10.1101/2022.11.16.22282431

**Authors:** Nefel Tellioglu, Rebecca H. Chisholm, Patricia Therese Campbell, Shelui Collinson, Joseph Timothy, Karsor Kollie, Samuel Zayzay, Angela Devine, Jodie McVernon, Michael Marks, Nicholas Geard

**Affiliations:** School of Computing and Information Systems, The University of Melbourne, Australia; Department of Mathematical and Physical Sciences, La Trobe University, Australia; Centre for Epidemiology and Biostatistics, Melbourne School of Population and Global Health, The University of Melbourne, Australia; Department of Infectious Diseases, University of Melbourne, at the Peter Doherty Institute for Infection and Immunity, Victoria, 3000, Australia; Centre for Epidemiology and Biostatistics, Melbourne School of Population and Global Health, The University of Melbourne, Melbourne, Australia; Clinical Research Department, Faculty of Infectious and Tropical Diseases, London School of Hygiene & Tropical Medicine, UK; Department of Disease Control, Faculty of Infectious and Tropical Diseases, London School of Hygiene & Tropical Medicine, UK; Ministry of Health, Liberia; Global and Tropical Health Division, Menzies School of Health Research, Charles Darwin University, Australia; Centre for Epidemiology and Biostatistics, Melbourne School of Population and Global Health, University of Melbourne, Australia; Department of Infectious Diseases, University of Melbourne, at the Peter Doherty Institute for Infection and Immunity, Melbourne, Australia; Victorian Infectious Diseases Reference Laboratory, The Royal Melbourne Hospital, at the Peter Doherty Institute for Infection and Immunity, Melbourne, Australia; Hospital for Tropical Diseases, University College London Hospital, London, UK; Division of Infection and Immunity, University College London, London, UK

**Keywords:** scabies, agent-based modelling, Liberia, mass drug administration, transmission

## Abstract

**Background:** Scabies is a parasitic infestation with high global burden. Mass drug administrations (MDAs) are recommended for communities with a scabies prevalence of >10%. Quantitative analyses are needed to demonstrate the likely effectiveness of MDA recommendations. In this study, we compare the effectiveness of differing MDA strategies, supported by improved treatment access, on scabies prevalence in Monrovia, Liberia.

**Methods:** We developed an agent-based model of scabies transmission calibrated to demographic and epidemiological data from Monrovia. We used this model to compare the effectiveness of MDA scenarios for achieving scabies elimination and reducing scabies burden, as measured by time until recrudescence following delivery of an MDA and disability-adjusted-life-years (DALYs) averted. We also investigated the additional impact of improving access to scabies treatment following delivery of an MDA.

**Results:** Our model showed that 3 rounds of MDA delivered at 6-month intervals and reaching 80% of the population could reduce prevalence below 2% for 3 years following the final round, before recrudescence. When MDAs were followed by increased treatment uptake, prevalence was maintained below 2% indefinitely. Increasing the number of and coverage of MDA rounds increased the probability of achieving elimination and the DALYs averted.

**Conclusions:** Our results suggest that acute reduction of scabies prevalence by MDA can support a transition to improved treatment access. This study demonstrates how modelling can be used to estimate the expected impact of MDAs by projecting future epidemiological dynamics and health gains under alternative scenarios.

**Summary:** We use an agent-based model to demonstrate that mass drug administration (MDA) programs can achieve sustained reduction in scabies prevalence. However, effective MDAs must be accompanied by systemic changes that increase the rate of scabies treatment to prevent recrudescence.

## Background

Scabies is a parasitic infestation commonly observed in tropical and resource-poor settings [1]. The primary manifestations of scabies are severe pruritic skin lesions on the host [2,3]. Scratching due to scabies infestation often leads to secondary bacterial infections such as Group A *Streptococcus* infections [4,5]. In 2016, it was estimated that scabies caused 3.8 million disability-adjusted life-years (DALYs) globally [6].

To reduce the scabies burden, an ivermectin-based mass drug administration (MDA) strategy is recommended when the community-level scabies prevalence is above 10% [7]. Several clinical studies in island populations have shown a significant reduction in scabies prevalence one year after a single round of MDA [8–10]; although one study did not demonstrate a reduction which was believed to be due to high human mobility [11]. Longer term follow-up has demonstrated some rebound in prevalence following cessation of MDA delivery [8,12,13]. To date, most MDA studies have taken place in isolated communities [8,10,11,14,15], making their direct application to larger areas and to highly interconnected urban settings uncertain.

Based on previous observations and practical considerations around programme delivery, the current recommendation is for 3 to 5 rounds of MDA applied at annual intervals [7]. Surveillance of the population should be continued for at least one year following the last MDA round to assess whether the prevalence has reached the target level of less than 2%. When observed prevalence is below this ‘stopping threshold’ it is proposed that annual MDA rounds can be stopped [2]. Following cessation of MDA, health systems need to be strengthened to ensure access to ongoing treatment of scabies for sustained control [7]. As there are no clear recommendations for communities with a prevalence between 2% and 10%, and existing recommendations are based on limited evidence and expert opinion, the World Health Organization (WHO) has highlighted the need for further modelling research to estimate the likely effectiveness of MDA strategies [7].

Previous modelling studies have estimated the likely impact of scabies interventions including MDAs [16–19], but have not explicitly investigated the current MDA recommendations. In this study, we use an agent-based model of scabies transmission, calibrated to demographic and epidemiologic data from Monrovia, to estimate the effectiveness of alternative recurring MDA strategies in an urban population with a starting prevalence of around 10%, and estimate their longer-term impact, independently and in combination with improved treatment access.

## Methods

### Demographic and epidemiologic model and data

A cross-sectional survey conducted in Monrovia, Liberia in 2020 found a community-level scabies prevalence of 9.3% [20]. Since the prevalence is in the range of 2–10%, it is not clear whether an MDA should be applied in Monrovia, and whether an MDA strategy would be effective to reduce the scabies burden. We extended an existing framework of disease transmission in an age- and household-structured population [21] (Supplementary Section S1). We calibrated demographic parameters so that the model generated household size and age distributions corresponding to those observed in Monrovia, Liberia [20] (Supplementary Section S2.1). We used a susceptible-infectious-susceptible (SIS) model to generate the dynamics of scabies infestation and transmission in this population. We assumed that clearance of a scabies infestation was primarily driven by treatment rather than natural recovery and that the duration of infestation therefore corresponds to the time until treatment [22]. As the average time until scabies treatment is not known for Liberia, we assumed a mean infestation duration of 90 days, based on health system visit data from Liberia [23]. We used Bayesian inference [24] to calibrate transmission coefficients to match age- and household size-specific patterns of scabies incidence observed in Monrovia [20] (Supplementary Section S2.2).

### MDA Scenarios

We defined MDA scenarios in terms of the number of rounds, time interval between rounds, population coverage (percentage of population receiving treatment), population selection (whether individuals or households are selected randomly), and treatment efficacy (the probability of recovery of an infested person following treatment) (Table 1). In the individual-based population selection method the participation of individuals was independent across each round e.g. an individual had the same probability of participating in rounds 1, 2 and 3. In the household-based selection we assumed that the participation of a household was independent across each round but that within each household every member either did or did not receive MDA. We assumed that each MDA round consists of two doses of ivermectin given at 7–14 days intervals, and that every selected individual in each round received two doses [7]. We simulated scenarios for all combinations of MDA parameters shown in Table 1.

**Table 1:** MDA parameters and their values.

| **Parameters** | **Values** | **Reference** |
| --- | --- | --- |
| Rounds of MDA | 3, 5 | [7] |
| Time interval between rounds | 1 year, 6 months, 2 years | [7] |
| Population coverage (% of target population receiving treatment) | 60%, 70%, 80%, 90% | 80% [8], 85% [25], 81% [11], >80% [7] |
| Population selection method | Random, Household-based | [7,26] |
| Treatment efficacy (after two doses) | 85%, 90%, 95%<sup>1</sup> | 94% [25], 90% [27], 88% [8] |

Notes:
<sup>1</sup> Results with 85% and 95% treatment efficacy are presented in Supplementary Section S6.1.

We compared the effectiveness of the selected MDA strategies in the absence and presence of importation (one imported case per week) (Supplementary Section S6.2). For each run, we recorded whether a prevalence of less than 2% was achieved one year after the last MDA round, whether scabies was eliminated (zero infestations), the number of undiscounted DALYs averted per 10,000 people over 20 years, and the time until prevalence recrudesced to its baseline (pre-MDA) level (Supplementary Section S4). As the simulation model is stochastic, we ran each scenario 100 times and calculated means and 95% quantiles. We used these values to calculate the probability of decreasing prevalence to below the proposed stopping threshold of 2%, the probability of achieving elimination, an estimation of DALYs averted, and the distribution of the time gained until another MDA strategy would be necessary (when the prevalence reaches pre-MDA levels).

### Systemic Changes

The long duration of infestation is a likely contributor to high scabies prevalence. This duration corresponds to the time until treatment as there is no natural recovery from scabies [22]. Access to health services and healthcare-seeking behaviour (normalisation) are factors that affect the time until treatment [28,29]. MDAs may provide an opportunity to decrease the average duration of infestation in several ways. By reducing prevalence, MDA strategies may help break the normalisation of scabies and encourage people to seek treatment earlier [28,29]. Second, a healthcare system will be better able to provide treatment for every affected person when there are fewer such individuals in society [2]. However, in order for MDAs to be followed by systemic changes, post-MDA drug treatment must be available in the primary health care system. We selected two MDA strategies (MDA strategies applied with random individual selection in 3 annual rounds and 60% and 80% population coverage) to understand the impact of systemic changes. For selected scenarios, we estimated the additional impact of longer-term changes to the health system by reducing the time it takes for someone to receive effective treatment for scabies by 10% and 20% after the last MDA round.

## Results

### The probability of decreasing scabies prevalence below the stopping threshold increases as the time interval between rounds is reduced

We observed that the probability of achieving a prevalence below 2% increases with a shorter time interval between MDA rounds (Figure 1).

**Figure 1:**
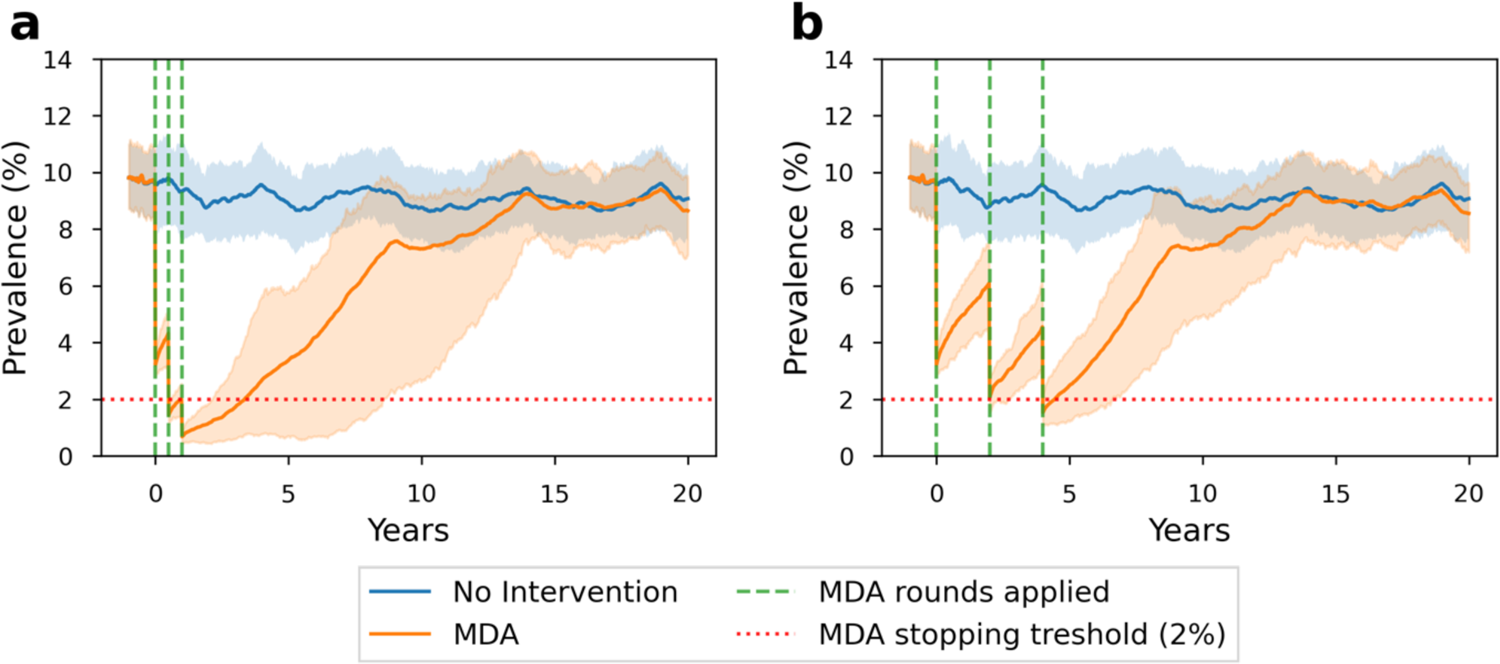
The scabies prevalence in 20 years with MDA strategies consisting of 3 rounds, 80% population coverage, random individual selection, and (a) six-month (b) two-year time intervals. The green dashed lines show when MDA rounds are applied. The red dotted lines represent MDA stopping threshold (2%). It is assumed that there is no scabies importation.

For an annual MDA strategy with 3 rounds, at least 80% population coverage was necessary to reach the stopping threshold in at least 75% of the simulations (Figure 2). At 90% coverage, prevalence less than the stopping threshold was achieved in more than 90 out of 100 runs, irrespective of number of rounds, interval between rounds and method of selection.

**Figure 2:**
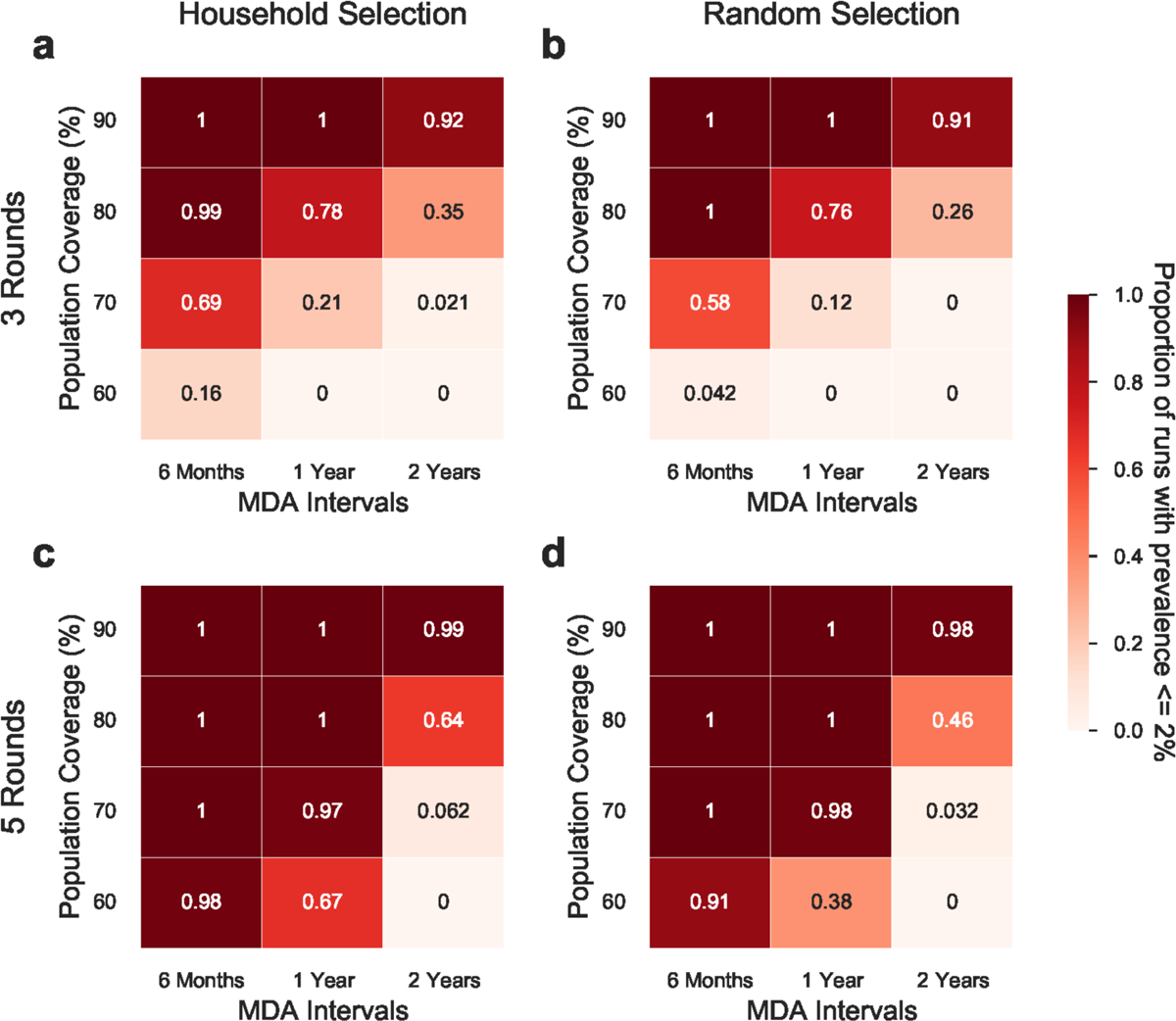
The proportion of simulation runs with less than 2% prevalence is achieved in differing MDA strategies. The first column (a & c) shows the MDA strategies with household-based selection and the second column (b & d) shows the MDA strategies with random individual selection. The first row (a & b) shows the MDA strategies with three rounds and the second row (c & d) shows the MDA strategies with 5 rounds. Each figure is grouped by the population coverage in MDAs and MDA intervals. Each value is calculated 1 year after the last MDA round from 100 simulation runs. In these scenarios, it is assumed that there is no scabies importation.

In contrast, at the lowest level of coverage tested (60%), this same result could only be achieved when five rounds of MDA were conducted at an interval of 6 months between rounds. Household-based selection almost always produced a higher proportion of simulations with less than 2% prevalence than random selection.

### Both the probability of scabies elimination and overall DALYs averted increase as the number of MDA rounds and population coverage are increased

We found that an MDA strategy consisting of 5 rounds with at least 90% population coverage and at most a one-year time interval is necessary to eliminate scabies (Table 2). We observed more DALYs averted with more MDA rounds and a higher population coverage (Table 2). The probability of scabies elimination one year after program cessation increased when we considered shorter time intervals between rounds, as previously described [19]. We also found that the average number of DALYs averted was higher when the time interval between MDA rounds was reduced. In addition, we did not observe a notable impact of the method of population selection on DALYs averted in 20 years.

**Table 2:** Percentage of simulations with <=2% prevalence 1 year after the last MDA round, percentage of simulations with scabies elimination, DALYs averted per 10,000 people (mean, 2.5 – 97.5 quantiles), and time until *prevalence reaches baseline are presented for* ***MDA strategies with at least 80% population coverage***. *Time until prevalence returns to baseline is calculated among the runs in which scabies is not eliminated. MDA strategies are ordered from best to worst in terms of DALYs averted*.

| MDA rounds | Population selection | Population coverage (%) | MDA Interval | The percentage of runs with $\leq 2\%$ prevalence 1 year after the last MDA round (%) | Percentage of runs with scabies elimination (%) | DALYs averted per 10,000 people in 20 years (Mean, 2.5 – 97.5 quantiles) | Time until prevalence returns to baseline (mean, 2.5 – 97.5 quantiles, in years) |
| --- | --- | --- | --- | --- | --- | --- | --- |
| 5 | Household | 90 | 6 months | 100 | 18 | 341.7 (161.7, 487.4) | 20+ (11.6, 20+) |
| 5 | Random | 90 | 1 year | 100 | 10 | 330.8 (204.2, 477.5) | 20+ (12.7, 20+) |
| 5 | Household | 90 | 1 year | 100 | 10 | 326.1 (191.5, 462.4) | 20+ (12.7, 20+) |
| 5 | Random | 90 | 6 months | 100 | 16 | 325.0 (164.2, 488.9) | 20+ (11.7, 20+) |
| 5 | Household | 90 | 2 years | 99 | 0 | 307.5 (210.1, 429.2) | 20+ (13.8, 20+) |
| 5 | Random | 90 | 2 years | 98 | 3 | 306.2 (202.0, 437.7) | 20+ (13.5, 20+) |
| 5 | Random | 80 | 6 months | 100 | 0 | 298.0 (144.7, 461.8) | 20+ (11.2, 20+) |
| 5 | Household | 80 | 6 months | 100 | 0 | 283.4 (158.1, 458.3) | 20+ (11.6, 20+) |
| 5 | Household | 80 | 1 year | 100 | 0 | 269.8 (165.5, 458.6) | 20+ (11.5, 20+) |
| 5 | Random | 80 | 1 year | 100 | 0 | 248.3 (150.0, 419.0) | 18.2 (11.7, 20+) |
| 3 | Household | 90 | 6 months | 100 | 0 | 218.2 (91.1, 459.0) | 18.4 (8.2, 20+) |
| 5 | Household | 80 | 2 years | 64 | 0 | 212.5 (156.6, 291.7) | 16.4 (12.9, 20+) |
| 3 | Random | 90 | 6 months | 100 | 0 | 210.6 (106.4, 450.2) | 17.7 (7.7, 20+) |
| 3 | Household | 90 | 1 year | 100 | 0 | 203.6 (112.3, 433.3) | 17.7 (8.1, 20+) |
| 5 | Random | 80 | 2 years | 46 | 0 | 197.1 (144.6, 290.2) | 15.9 (12.4, 20+) |
| 3 | Random | 90 | 1 year | 100 | 0 | 188.6 (108.6, 441.9) | 13.6 (8.1, 20+) |
| 3 | Random | 90 | 2 years | 91 | 0 | 168.5 (108.5, 367.4) | 13.2 (8.5, 20+) |
| 3 | Household | 90 | 2 years | 92 | 0 | 159.3 (108.0, 243.6) | 12.8 (8.7, 17.8) |
| 3 | Household | 80 | 6 months | 99 | 0 | 143.9 (91.0, 267.5) | 12.3 (6.9, 17.9) |
| 3 | Random | 80 | 6 months | 100 | 0 | 134.3 (81.5, 237.0) | 12.2 (6.8, 18.4) |
| 3 | Household | 80 | 1 year | 78 | 0 | 127.2 (80.4, 208.8) | 11.8 (7.1, 13.8) |
| 3 | Random | 80 | 1 year | 76 | 0 | 127.1 (76.1, 184.2) | 11.2 (7.2, 20+) |
| 3 | Household | 80 | 2 years | 35 | 0 | 121.3 (80.1, 175.7) | 11.4 (8.0, 19.0) |
| 3 | Random | 80 | 2 years | 26 | 0 | 117.6 (82.6, 174.9) | 11.4 (7.6, 17.9) |

### Systemic changes coupled with MDA strategies can help us sustainably maintain the scabies prevalence at a lower level

We observed that reducing the time to routine treatment of scabies by 20% after the final MDA round results in the elimination of scabies in 36% of simulations, compared to 0% of simulations in scenarios with no reduction in the time to treatment, suggesting that, following MDA, health system or behavioural changes which result in a reduction in the infestation duration can sustainably maintain scabies prevalence at a lower level (Figure 3). These sustained reductions in prevalence post MDA occur because the shortened duration of infestation corresponds to reductions in the basic reproduction number, R0, from 1.24 (baseline) to 1.10 with a 10% reduction, and to 0.99 with a 20% reduction (Supplementary Section S3).

**Figure 3:**
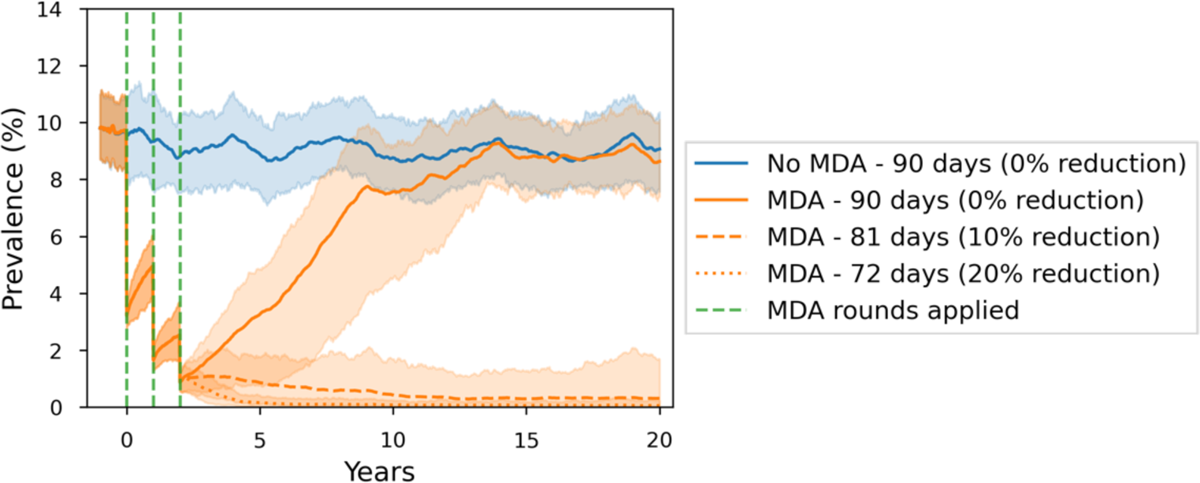
Scabies prevalence over 20 years with (orange) and without (blue) an MDA strategy. Green dashed lines show when MDA rounds are applied (years 0, 1, and 2). Solid lines represent scenarios with an average of 90 days (baseline – 0% reduction) duration of infestation throughout the simulation while dashed and dotted lines represent an average of 81 (10% reduction) and 72 (20% reduction) days duration of infestation after year 2, respectively. MDA strategies were applied with random individual selection in 3 annual rounds and 80% population coverage. There was no scabies importation.

## Discussion

In this study, we evaluated the effectiveness of various MDA strategies for reducing the scabies prevalence using an agent-based model calibrated to survey data collected in Monrovia, Liberia [20]. We observed that while MDAs can have short- and medium-term success [8,9,30,31] it remains likely that prevalence will return to pre-MDA levels over longer time periods, especially in the absence of health system or systemic behavioural changes [19]. However, sustainable improvements to scabies control can be achieved when MDAs are coupled with systemic changes that reduce the time it takes for scabies infestations to be successfully treated (Figure 3). When post-MDA treatment is available in the healthcare system, MDAs may naturally provide an environment for systemic changes by breaking the normalisation cycle and increasing the per capita rate of healthcare access of individuals with scabies [28,29] which highlights the importance of post-MDA treatment accessibility. Moreover, MDAs are likely to have a higher chance of sustainably maintaining scabies prevalence at a low level when supported by other interventions such as improved education and health system accessibility [1,2,32,33].

The current recommendation is for 3-5 rounds of MDA, stopping when prevalence is reduced below 2% [2,7]. We observed that prevalence can be reliably reduced below 2% provided that the population coverage is sufficient (Figure 2). We also observed that it is unlikely that MDAs will be sufficient to eliminate scabies without broader health system changes that reduce delays in treatment of scabies beyond the MDA setting (Figure 3).

Several existing studies use modelling to compare the effectiveness of scabies interventions including MDAs [16–19]. However, to our knowledge, this is the first modelling analysis to estimate the efficacy of recommended MDA strategies, as well as the first to quantify the role of community transmission in a sub-Saharan African setting. For instance, our model showed that among MDA strategies with >80% coverage, the DALYs averted per 10,000 people is increased from 120 to 340 years when the population coverage and number of rounds are increased. Our study provides a flexible modelling framework for scabies that can be calibrated to other settings to compare community-level interventions using DALYs and incidence statistics.

As with any modelling study, our results depend on assumptions made and are subject to some limitations. First, limited data were available to calibrate some model parameters, including duration of infestation and importation rate. Our sensitivity analyses established that estimates of MDA efficacy were sensitive to these parameters. Collection of additional data on time to treatment of scabies and population mobility may reduce the uncertainty around model estimates. Second, in this study, we report the ‘true’ prevalence of scabies in the modelled population. In reality, estimates of prevalence will depend on the sampling strategy used to detect infestations [34]. Monitoring the impact of MDAs and responding appropriately will thus require effective surveillance in addition to effective interventions. Third, in the population selection methods, we assumed that MDA rounds were independent from each other and every selected individual in each round received two doses of treatment within 7–14 days. Under these assumptions, we did not consider the impact of (1) non-compliance, where people receive no treatment or only one dose rather than two doses in each round [35,36], and (2) systematic non-treatment, where individuals receiving treatment in previous rounds are not treated in the following rounds [37]. Further sensitivity analyses could be performed to understand the impact of systematic non-treatment and non-compliance. Finally, we estimated DALYs averted under various MDA scenarios but did not consider the differing costs associated with each scenario. However, strategies that were likely to avert most DALYs were also likely to be the most resource intensive. The results presented here could form the basis of future cost-effectiveness analyses to provide a more robust basis for comparison [8,38].

In summary, our modelling study suggests that MDAs can play a critical role in scabies control by reducing the prevalence and maintaining it at a low level when combined with systemic changes. This study demonstrates how modelling can be used to refine and define effective interventions in reducing scabies burden in large populations with endemic prevalence.

## Data Availability

All data produced in the present study are available upon reasonable request to the authors

