## Supplementary Materials for "Evaluation of the effectiveness of mass drug administration strategies for reducing scabies burden in Monrovia, Liberia: An agent-based modelling approach"

### Supplementary material

Nefel Tellioglu
School of Computing and Information Systems, The University of Melbourne, Australia

Rebecca H. Chisholm
Department of Mathematical and Physical Sciences, La Trobe University, Australia
Centre for Epidemiology and Biostatistics, Melbourne School of Population and Global Health, The University of Melbourne, Australia

Patricia Therese Campbell
Department of Infectious Diseases, University of Melbourne, at the Peter Doherty Institute for Infection and Immunity, Victoria, 3000, Australia
Centre for Epidemiology and Biostatistics, Melbourne School of Population and Global Health, The University of Melbourne, Melbourne, Australia

Shelui Collinson
Clinical Research Department, Faculty of Infectious and Tropical Diseases, London School of Hygiene & Tropical Medicine, UK

Joseph Timothy
Department of Disease Control, Faculty of Infectious and Tropical Diseases, London School of Hygiene & Tropical Medicine, UK

Karsor Kollie
Ministry of Health, Liberia

Samuel Zayzay
Ministry of Health, Liberia

Angela Devine
Global and Tropical Health Division, Menzies School of Health Research, Charles Darwin University, Australia
Centre for Epidemiology and Biostatistics, Melbourne School of Population and Global Health, University of Melbourne, Australia

Jodie McVernon
Department of Infectious Diseases, University of Melbourne, at the Peter Doherty Institute for Infection and Immunity, Melbourne, Australia
Victorian Infectious Diseases Reference Laboratory, The Royal Melbourne Hospital, at the Peter Doherty Institute for Infection and Immunity, Melbourne, Australia

Michael Marks
Clinical Research Department, Faculty of Infectious and Tropical Diseases, London School of Hygiene & Tropical Medicine, UK
Hospital for Tropical Diseases, University College London Hospital, London, UK
Division of Infection and Immunity, University College London, London, UK

Nicholas Geard *
School of Computing and Information Systems, The University of Melbourne, Australia

Figure S9: (a) Mean and 2.5–97.5% quantiles of overall prevalence, (b) mean and 2.5–97.5% quantiles of prevalence in age groups, (c) mean and 2.5–97.5% quantiles of prevalence in

Figure S12: The proportion of simulation runs with less than 2% prevalence is achieved in differing MDA strategies **with an average of duration of infestation of 150 days**. The first column (a & c) are the MDA strategies with household-based selection and the second column (b & d) are the MDA strategies with random individual selection. The first row (a & b) are the MDA strategies with three rounds and the second row (c & d) are the MDA strategies with 5 rounds. Each panel is grouped by the population coverage in MDAs and MDA intervals. Each value is calculated from 100 simulation runs. In these scenarios, it is assumed that there is no scabies importation...........................................................................16
Figure S13: Mean and 2.5–97.5 quantiles of DALYs averted per 10000 people in differing MDA strategies **with an average duration of infestation of 150 days**. The values are calculated in 20 years starting from the first MDA round. The first column (a & c) are the MDA strategies with household-based selection and the second column (b & d) are the MDA strategies with random individual selection. The first row (a & b) are the MDA strategies with three rounds and the second row (c & d) are the MDA strategies with 5 rounds. Each panel is grouped by the population coverage in MDAs and MDA intervals. Each value is calculated from 100 simulation runs. In these scenarios, it is assumed that there is no scabies importation.......................................................................................................................17
Figure S14: Mean and 2.5–97.5 quantiles of DALYs averted per 10000 people in differing MDA strategies with **an average duration of infestation of 90 days**. The values are calculated in 20 years starting from the first MDA round. The first column (a & c) are the MDA strategies with household-based selection and the second column (b & d) are the MDA strategies with random individual selection. The first row (a & b) are the MDA strategies with three rounds and the second row (c & d) are the MDA strategies with 5 rounds. Each panel is grouped by the population coverage in MDAs and MDA intervals. Each value is calculated from 100 simulation runs. In these scenarios, it is assumed that there is no scabies importation.......................................................................................................................19
Figure S15: The proportion of simulation runs with a prevalence of less than 2% is achieved in differing MDA strategies having a treatment efficacy of 85% (rather than 90%) in scenarios with **an average duration of infestation of 90 days**. The first column (a & c) are the MDA strategies with household-based selection and the second column (b & d) are the MDA strategies with random individual selection. The first row (a & b) are the MDA strategies with three rounds and the second row (c & d) are the MDA strategies with 5 rounds. Each panel is grouped by the population coverage in MDAs and MDA intervals. Each value is calculated from 100 simulation runs. It is assumed that there is no scabies importation.......................................................................................................................22

Figure S16: The proportion of simulation runs with a prevalence of less than or equal to 2% is achieved in differing MDA strategies having a treatment efficacy of 95% (rather than 90%) in scenarios with **an average duration of infestation of 90 days**. The first column (a & c) are the MDA strategies with household-based selection and the second column (b & d) are the MDA strategies with random individual selection. The first row (a & b) are the MDA strategies with three rounds and the second row (c & d) are the MDA strategies with 5 rounds. Each panel is grouped by the population coverage in MDAs and MDA intervals. Each value is calculated from 100 simulation runs. ............................................................25
Figure S17: The scabies prevalence in 20 years with (orange) and without (blue) MDA strategy in scenarios (a) 60% population coverage – no importation, (b) 80% population coverage – no importation, (c) 60% population coverage – with scabies importation, and (d) 80% population coverage – with scabies importation **with an average infestation duration of 90 days**. The green lines show when MDA rounds are applied (years 0, 1, and 2). Solid lines show scenarios with an average of 90 days (baseline – 0% reduction) duration of infestation throughout the simulation while dashed and dotted lines show an average of 81 (10% reduction) and 72 (20% reduction) days duration of infestation after year 2, respectively. MDA strategies are applied with random individual selection in 3 annual rounds. In scenarios with scabies importation, there is only one weekly scabies importation. Panel b represents results from Figure 3 in the main manuscript. .........................................28
Figure S18: The scabies prevalence in 20 years with (orange) and without (blue) MDA strategy in scenarios (a) 60% population coverage – no importation, (b) 80% population coverage – no importation, (c) 60% population coverage – with scabies importation, and (d) 80% population coverage – with scabies importation **with an average infestation duration of 150 days**. The green lines show when MDA rounds are applied (years 0, 1, and 2). Solid lines show scenarios with an average of 150 days (baseline) of the duration of infestation throughout the simulation while dashed and dotted lines show an average of 135 (10% reduction) and 120 (20% reduction) days of the duration of infestation after year 2, respectively MDA strategies are applied with random individual selection in 3 annual rounds. In scenarios with scabies importation, there is only one weekly scabies importation............29

**List of Tables**

Table S1: Population model parameters ......................................................................................8
Table S2: Estimation of $R_0$ in baseline and other scenarios with lower durations of infestation. $R_0$ estimations calculated using baseline duration of infestation and reduced duration of infestation are presented for simulations with average of 90 days and 150 days duration of infestation. .........................................................................................................13
Table S3: Percentage of simulations with <=2% prevalence 1 year after the last MDA round, percentage of simulations with scabies elimination, DALYs averted per 10,000 people (mean, 2.5–97.5 quantiles), and time until prevalence returns to baseline are presented in **MDA strategies with 3 rounds with an average duration of infestation of 150 days**. Time until prevalence returns to baseline is calculated among the runs in which scabies is not eliminated. It is assumed that there is no scabies importation. MDA strategies are ordered from best to worst in terms of DALYs averted. ............................................................................17
Table S4: Percentage of simulations with <=2% prevalence 1 year after the last MDA round, percentage of simulations with scabies elimination, DALYs averted per 10,000 people (mean, 2.5–97.5 quantiles), and time until prevalence returns to baseline are presented in **MDA strategies with 5 rounds with an average of duration of infestation of 150 days.** Time until prevalence returns to baseline is calculated among the runs in which scabies is not

eliminated. It is assumed that there is no scabies importation. MDA strategies are ordered from best to worst in terms of DALYs averted......18
Table S5: Percentage of simulations with <=2% prevalence 1 year after the last MDA round, percentage of simulations with scabies elimination, DALYs averted per 10,000 people (mean, 2.5–97.5 quantiles), and time until prevalence returns to baseline are presented in **MDA strategies with 3 rounds in scenarios with an average duration of infestation of 90 days**. Time until prevalence returns to baseline is calculated among the runs in which scabies is not eliminated. It is assumed that there is no scabies importation. MDA strategies are ordered from best to worst in terms of DALYs averted......20
Table S6: Percentage of simulations with <=2% prevalence 1 year after the last MDA round, percentage of simulations with scabies elimination, DALYs averted per 10,000 people (mean, 2.5–97.5 quantiles), and time until prevalence returns to baseline are presented in **MDA strategies with 5 rounds in scenarios with an average duration of infestation of 90 days**. Time until prevalence returns to baseline is calculated among the runs in which scabies is not eliminated. It is assumed that there is no scabies importation. MDA strategies are ordered from best to worst in terms of DALYs averted......21
Table S7: Percentage of simulations with <=2% prevalence 1 year after the last MDA round, percentage of simulations with scabies elimination, DALYs averted per 10,000 people (mean, 2.5–97.5 quantiles), and time until prevalence returns to baseline are presented in **MDA strategies with 3 rounds and 85% treatment efficacy** in scenarios with **an average duration of infestation of 90 days.** Time until prevalence returns to baseline is calculated among the runs in which scabies is not eliminated. It is assumed that there is no scabies importation. MDA strategies are ordered from best to worst in terms of DALYs averted. ....22
Table S8: Percentage of simulations with <=2% prevalence 1 year after the last MDA round, percentage of simulations with scabies elimination, DALYs averted per 10,000 people (mean, 2.5 – 97.5 quantiles), and time until prevalence returns to baseline are presented in **MDA strategies with 5 rounds and 85% treatment efficacy** in scenarios with **an average duration of infestation of 90 days.** Time until prevalence returns to baseline is calculated among the runs in which scabies is not eliminated. It is assumed that there is no scabies importation. MDA strategies are ordered from best to worst in terms of DALYs averted. ....23
Table S9: Percentage of simulations with <=2% prevalence 1 year after the last MDA round, percentage of simulations with scabies elimination, DALYs averted per 10,000 people (mean, 2.5–97.5 quantiles), and time until prevalence returns to baseline are presented in **MDA strategies with 3 rounds and 95% treatment efficacy** in scenarios with **an average duration of infestation of 90 days.** Time until prevalence returns to baseline is calculated among the runs in which scabies is not eliminated. It is assumed that there is no scabies importation. MDA strategies are ordered from best to worst in terms of DALYs averted. ....25
Table S10: Percentage of simulations with <=2% prevalence 1 year after the last MDA round, percentage of simulations with scabies elimination, DALYs averted per 10,000 people (mean, 2.5 – 97.5 quantiles), and time until prevalence returns to baseline are presented in **MDA strategies with 5 rounds and 95% treatment efficacy** in scenarios with **an average duration of infestation of 90 days.** Time until prevalence returns to baseline is calculated among the runs in which scabies is not eliminated. It is assumed that there is no scabies importation. MDA strategies are ordered from best to worst in terms of DALYs averted. ....26

### S1. Agent-based model

We use the disease transmission model with age and household structure from Geard, et al [1]. In this model, a synthetic population is generated with given demographic parameters. Then, disease transmission is simulated in the generated synthetic population. Using this framework, disease transmission can be simulated over an extended period of time (e.g., decades) with plausible age and household dynamics.

We modelled scabies transmission using an SIS model, where infectious individuals return to the susceptible class on recovery [2]. The infectious duration is sampled from an Erlang distribution (with shape parameter k = 3) and a given mean infectious duration. Two types of transmission are simulated in the model: household and community transmission. In addition, there is an external exposure rate that simulates people becoming infected from a source outside of the model population. The model assumes that a person makes contact with all other members of their household in every time step and that they make contact with other people in the community at age-specific rates. These age-specific community contact rates are represented as a contact matrix which is calculated by the activity levels of age groups. The activity levels are given as the difference between the "all contacts" matrix and the "household contacts" matrix for Liberia by Prem, et al [3] since household contacts are represented explicitly in the model.

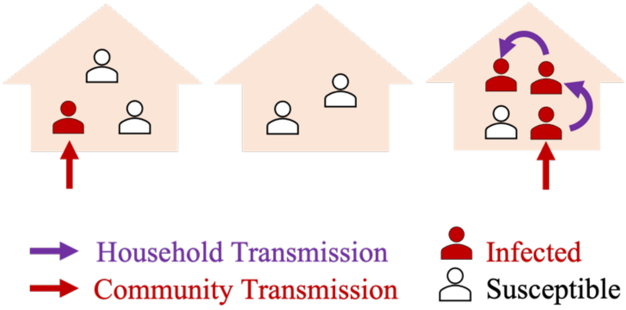

*Figure S1: Illustration of community and household transmission in our model. The source (infector) in a community transmission is unknown but the source in a household transmission is known.*

The probability of a person becoming infected in each time step is given by:

$$Probability\ of\ infection = 1 - e^{-foi_{iNH}}$$

where *foi* is the force of infection on an individual in age class *i*, belonging to a household of size $N_H$ [1] The force of infection *foi* is made up of components related to household and community transmission:

$$foi_{iNH} = \underbrace{q_h * \frac{I_H}{(N_H - 1)}}_{\text{Household Transmission}} + \underbrace{q_C * \sum_{j=1} \eta_{ij} \frac{I_j}{N_j}}_{\text{Community Transmission}}$$

where:

- $q_h$ = Household transmission coefficient
- $q_C$ = Community transmission coefficient

- $\eta_{ij}$ = Number of daily community contacts between age groups $i$ and $j$
- $I_H$ = Number of infected individuals in a household
- $I_j$ = Number of infected individuals in age group $j$

In addition to the endemic transmission, there is also a chance of susceptible individuals being infected by external sources with a probability of external exposure rate multiplied by population size. The rate of household transmission depends on the proportion of infectious people in the household. The rate of community transmission depends on the age-dependent community contact rates $\eta_{ij}$ and the proportion of infected people in each age class.

### S2. Calibration of the agent-based model

A cross-sectional survey conducted in Monrovia, Liberia in 2020 found a community-level scabies prevalence of 9.3% [4]. While the survey provides a detailed picture of scabies prevalence in sub-Saharan Africa, it is cross-sectional and hence provides only limited information about transmission dynamics and the likely efficacy of control strategies. Since the prevalence is in the range of 2−10%, it is not clear whether an MDA should be applied in Monrovia, and whether an MDA strategy would be effective to reduce the scabies burden.

We first calibrated the population model to the demographic characteristics of the peri-urban settings of Liberia (Section S2.1). Then, we calibrated the disease dynamics to the survey data (Section S2.2).

#### S2.1. Calibration of the population model

We first calibrated the population demography in our model to match that of the study population [5] and compared characteristics of age and household observed in the synthetic population with the survey data from Monrovia [4] (Table S1). We run the population model for 100 years starting from an initial population size of 5000. The population size became ~13600 after the 100-year burn-in period.

However, the Liberia population characteristics were different from the survey data (Figure S2). Therefore, we decided to use the death rates [6] and fertility rates [7] of Zambia as these datasets provided a better representation of household- and age-specific characteristics of the survey data collected in New Kru Town, Monrovia [4]. The age and household-size distributions of the synthetic population and original dataset are shown in Figure S3 and Figure S4. The synthetic population in our model represents the demographic characteristics of the survey data.

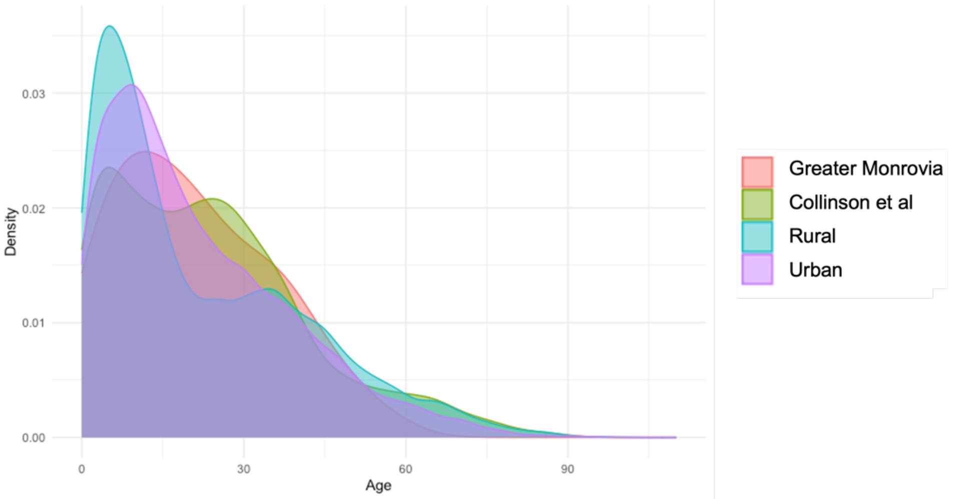

*Figure S2: Comparison of age distribution in survey data collected in New Kru Town, Monrovia (Collinson et al - green)* [4] *and age distributions for Greater Monrovia (red), rural (blue), urban (purple) taken from the World Bank data* [5].

*Table S1: Population model parameters*

| **Parameter** | **Description** | **Values** |
|---|---|---|
| Initial population size | Number of individuals in population at start of simulation. | 5000 |
| Population growth rate | Annual rate of change in population size due to natural increase. | 0.01 |
| Immigration rate | Annual rate of change in population size due to immigration. | 0.0 |
| Age- and sex-specific mortality probabilities | Annual probabilities of death by sex and year of age. | Dataset |
| Age-specific relative fertility probabilities | Relative probabilities, given the birth of a child, that the mother is of a specified age. | Dataset |
| Birth gap (mean and SD) | Parameters governing the minimum inter-birth interval. | (270, 0) |
| Couple formation parameters | Age range that a currently single individual is eligible to form a couple, and annual probability that this will occur. | (15, 60), 0.039 |
| Partner age difference (mean and SD) | Parameters governing the sex-dependent age difference between partners during couple formation. | (2, 2) |
| Couple dissolution parameters | Age range that a currently coupled individual is eligible to dissolve that couple, and annual probability that this will occur. | (18, 60), 0.001 |
| Leaving home parameters | Minimum age at which an individual currently living with a parent/guardian will form a new single-person household, and annual probability that this will occur. | 18, 0.008 |

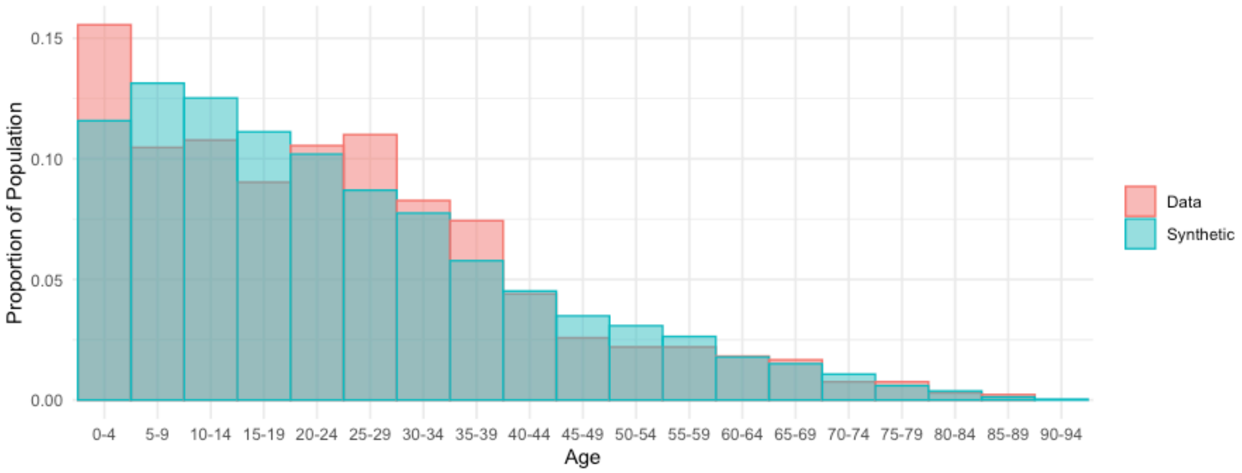

*Figure S3: Age Distribution of the Synthetic Population (blue) and Monrovia Dataset (red) at year 0 after 100-year burn-in period*

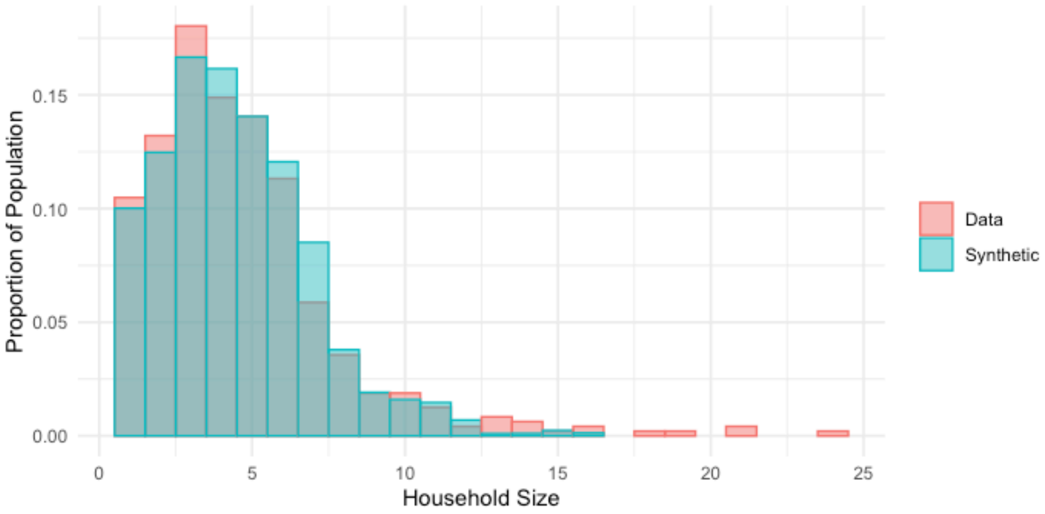

*Figure S4: Household Size Distribution of the Synthetic Population (blue) and Monrovia Dataset (red) at year 0 after 100-year burn-in period*

### S2.2. Calibration of the disease model

This section summarises the calibration of our age and household structured model to data from the household scabies survey conducted in Monrovia, Liberia. We calibrated household and community transmission coefficients to match the age and household size specific patterns of scabies incidence. We applied The Bayesian Optimization in Likelihood-Free Inference (BOLFI) framework to calibrate the scabies model to the Liberian population [8]. To begin, we assumed a mean duration of infestation of 90 days which represents the time until seeking treatment. We used BOLFI to calibrate two unknown parameters: household and community transmission coefficients ($q_h$ and $q$). Using BOLFI involves defining a set of summary statistics that can be calculated from both simulated and real data, and searching the ($q_h$, $q$) parameter space for regions that minimise the distance between the summary statistics from the simulated and real data.

We used 33 summary statistics (captured at endemic equilibrium) in the calibration process:

-overall prevalence in the population (with weight 1),
-prevalence in 16 age groups (with weights as the percentage of people in the given population groups),
-prevalence in 12 household size groups (with weights as the percentage of people in the given population groups),
-percentage of households with zero, at least one, at least two, at least three cases (with weights as 0.25 summing to 1).

These statistics are collected from a new generated sample quarterly for five years (20 times total) in every simulation run. To match the sample characteristics of the data, a sample with size 1300 is selected uniformly at random from 480 houses selected uniformly at random. We used logarithm of Euclidian distance as the distance function.

The result of the model calibration is shown in Figure S5. In this plot, darker regions indicate a greater belief in the true values of the parameters $q_h$ and $q$. We simulated the model using 100 values of the $q$ & $q_h$ parameters generated from the posterior distribution (red points in Figure S6). The overall prevalence, prevalence in age and household groups are compared with the real data in Figure S7.

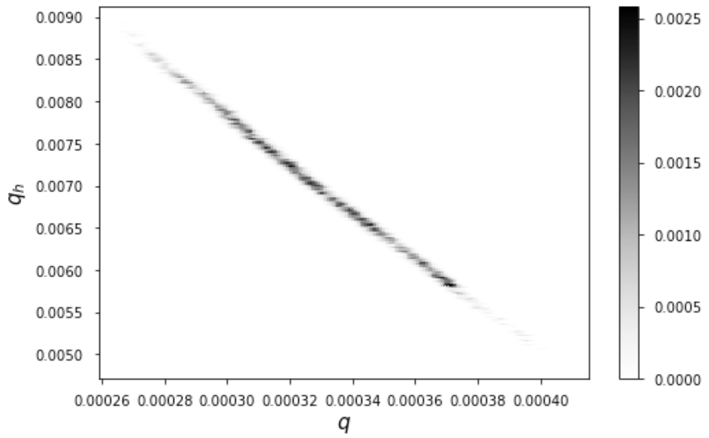

*Figure S5: Marginal posterior distribution for the household and community transmission coefficients ($q_h$ and $q$) estimated using BOLFI*

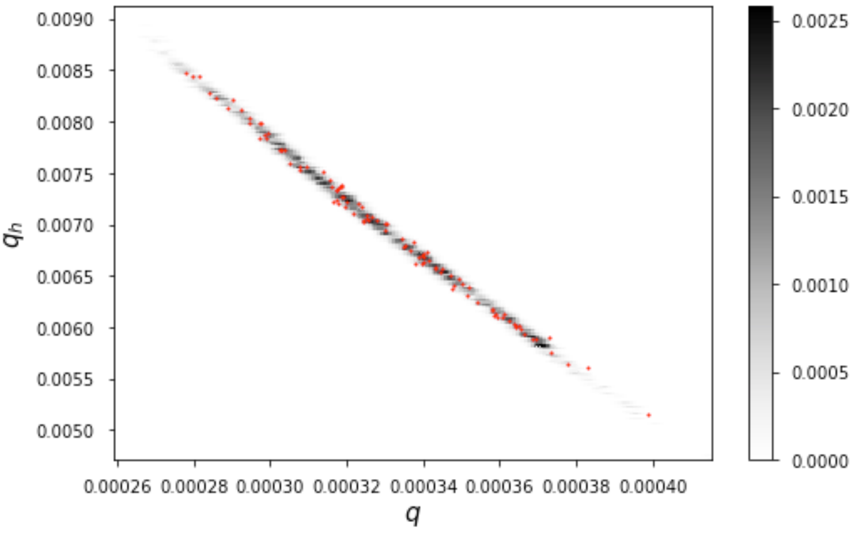

*Figure S6: Marginal posterior distribution for the household and community transmission coefficients ($q_h$ and q) and sampled points from posterior (red)*

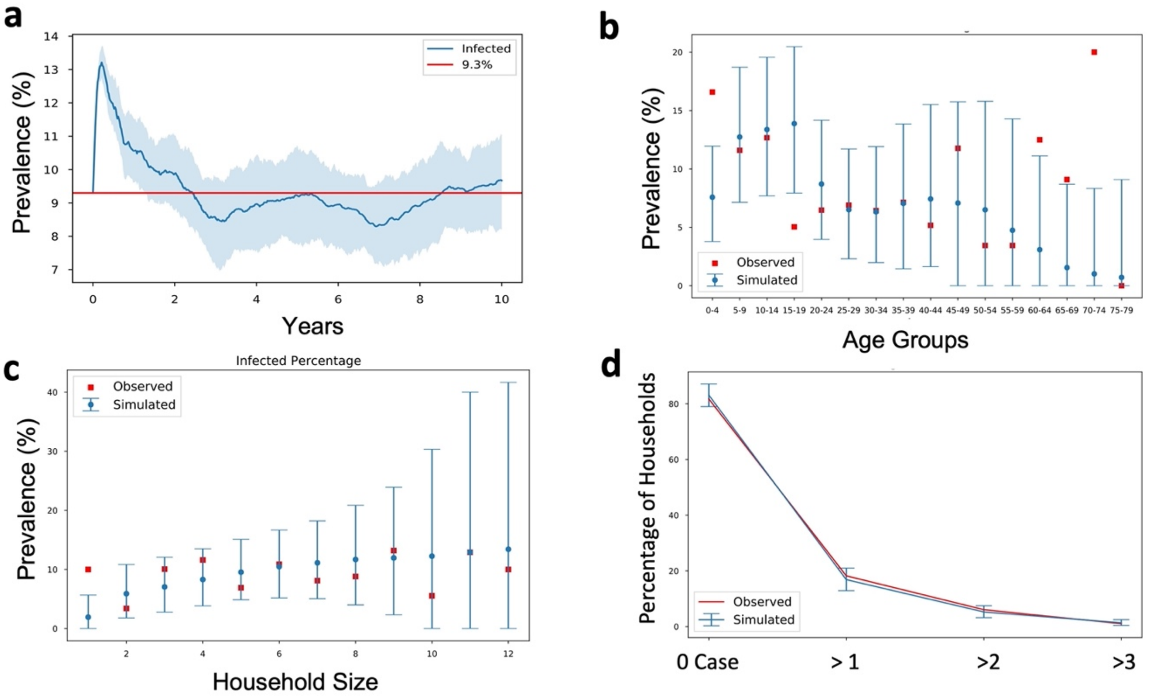

*Figure S7: (a) Mean and 2.5–97.5% quantiles of overall prevalence, (b) mean and 2.5–97.5% quantiles of prevalence in age groups, (c) mean and 2.5–97.5% quantiles of prevalence in household groups, (d) mean and 2.5–97.5% quantiles of the percentage of household groups with zero, at least one, at least two, at least three cases. In panels, blue represents simulation results and red represents the survey data. Data is captured quarterly after year 5 in panes (b), (c), and (d).*

Using the calibrated model, we next looked at the community transmission percentage, i.e., the percentage of all transmission events which occur outside of households. The calibrated model shows that community transmission is responsible for an average of 50% of cases with 2.5–97.5% quantiles of [42%, 59%] of all the community and household transmissions (Figure S8). While the average community transmission percentage of people between ages of 5−19 is 60%, the percentage decreases in older age groups suggesting that young people are more likely to introduce scabies into households. The percentage of infestations acquired in households increases from 0% to 50% as household size increases from one to five members, stabilising at 50% thereafter, suggesting that larger households do not necessarily have higher household transmission. Note that quantiles are very wide among older age groups and very large households as these occur less commonly in the simulated populations. We observed that the community transmission percentage was quite sensitive to the precise combination of $q$ and $q_h$ values used.

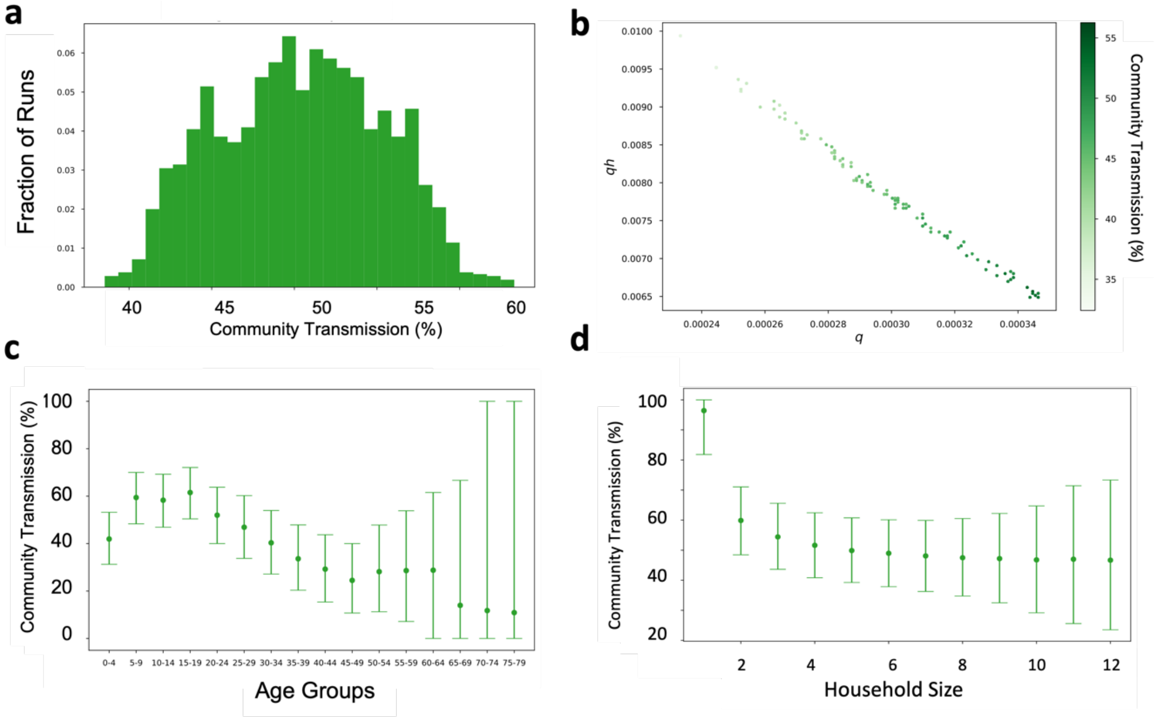

*Figure S8: (a) Histogram of overall community transmission percentage, (b) distribution of overall community transmission percentage, (c) mean and 2.5–97.5% quantiles of community transmission percentages in age groups and (d) mean and 2.5–97.5% quantiles of community transmission percentages in household size groups. Data is captured quarterly after year 5 in 100 simulations.*

### S3. $R_0$ Estimation

We have estimated $R_0$ by running simulations. We initialized the simulation with one person having scabies and simulated the epidemic for 2 years (5 years) for scenarios with 90 days (150 days) duration of infestation. We run the simulation 10000 times (1000 times for each combination of $q$ and $q_h$ parameters).

As only the source ID of household transmission is recorded in our modelling framework, we first estimated the secondary household infestations from the first person infested in every household in the first year (Algorithm 1). Then we estimated the secondary infections rate by dividing secondary household infections rate by household transmission percentage and we estimated $R_0$ by taking the mean of all the secondary infections rate.

*Algorithm 1: Pseudo code of $R_0$ estimation*

```
secondary_hh_infections_list = []

for every simulation run:
   if no transmission occurred:
      Add 0 to secondary_hh_infections_list
   else:
      if there is at least one community transmission:
         if the first transmission is in the household:
             Add the number of household infections from the first infected seed
            for every other person, agent i infected in the community and brought
       scabies to    their households for the first time:
                Add the number of household infections from agent i
      else: #there is only hh transmission
          Add the number of household infections from the first infected seed in
       the first year
Calculate household transmission percentage among all the transmission events
Calculate all secondary infections by dividing every item in
secondary_hh_infections_list by household transmission percentage
Calculate the mean and quantiles of all secondary infections
```

We estimated $R_0$ in scenarios with baseline duration of infestation and reduced duration of infestations with average of 90 days and 150 days (Table S2).

*Table S2: Estimation of $R_0$ in baseline and other scenarios with lower durations of infestation. $R_0$ estimations calculated using baseline duration of infestation and reduced duration of infestation are presented for simulations with average of 90 days and 150 days duration of infestation.*

| | **Baseline** | | **10% Reduction** | | **20% Reduction** | |
|---|---|---|---|---|---|---|
| | **90 days** | **150 days** | **81 days** | **135 days** | **72 days** | **120 days** |
| **$R_0$** | 1.24 | 1.237 | 1.107 | 1.18 | 0.989 | 1.04 |

### S4. Estimation of disability-adjusted life-years (DALYs)

We summed the duration of infestation for every infested person in 20 years to estimate the total person-years with scabies for 20 years starting from the beginning of the first MDA round. After calculating the mean and quantiles of the total person-years, we estimated the mean and quantiles of DALYs by multiplying the mean and quantiles of the total person-years by the disability weight assigned to scabies, 0.027 [9]. We then divided DALYs estimations for the entire population by "the average population size in 20 years/10,000" to estimate DALYs per 10,000 people.

In order to calculate DALYs averted, we estimated DALYs for every run for the "No MDA" scenario. Then, we subtracted DALYs of MDA scenarios from DALYs of "No MDA" scenarios for each parameter set. Then, we calculated DALYs averted by taking the mean and quantiles of obtained results.

### S5. Sensitivity analysis of duration of infestation

We conducted a sensitivity analysis around the average infestation days. We set the average duration of infestation as 150 days rather than 90 days. We first calibrated the parameter set ($q$ and $q_h$) using 150 days by using BOLFI. We ended up with quite similar age- and household-specific scabies prevalence in the calibrated model (Figure S9 and Figure S10) as model calibration decreased $q$ and $q_h$ values in accordance with the increase in the duration of infestation (Figure S10.b).

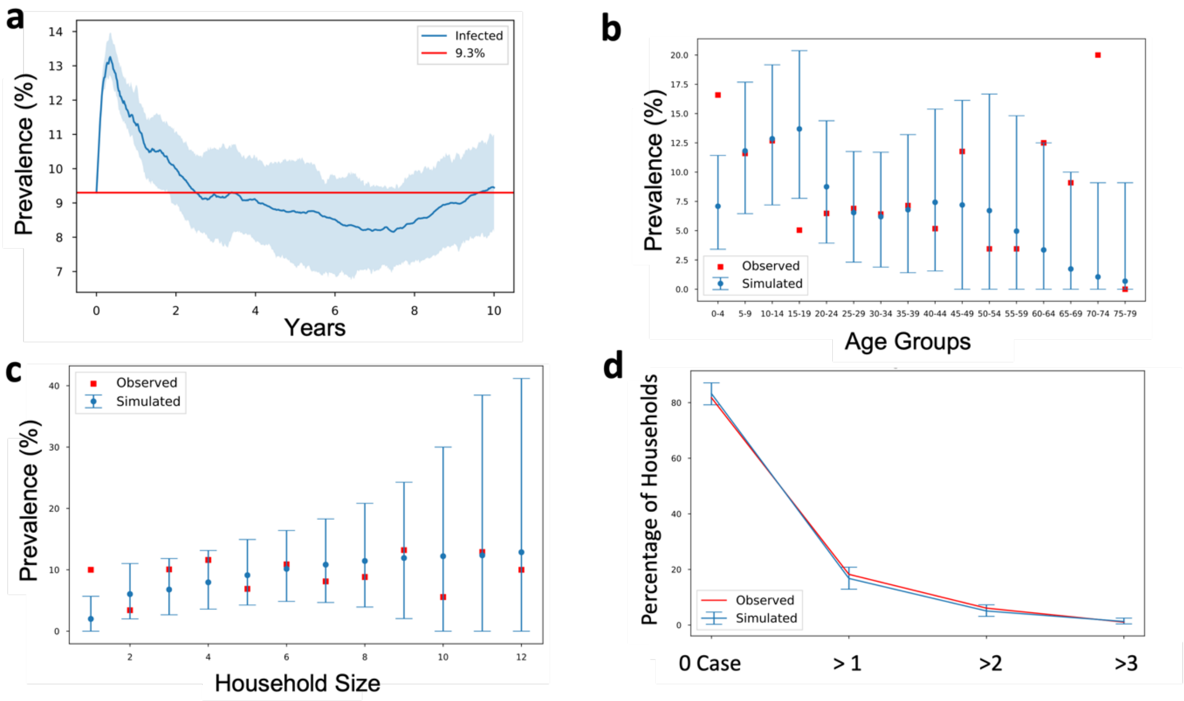

*Figure S9: (a) Mean and 2.5–97.5% quantiles of overall prevalence, (b) mean and 2.5–97.5% quantiles of prevalence in age groups, (c) mean and 2.5–97.5% quantiles of prevalence in household groups, (d) mean and 2.5–97.5% quantiles of the percentage of household groups with zero, at least one, at least two, at least three cases **with an average of 150 days of infestation duration**. In panels, blue represents simulation results and red represents the survey data. Data is captured quarterly after year 5 in panels (b), (c), and (d).*

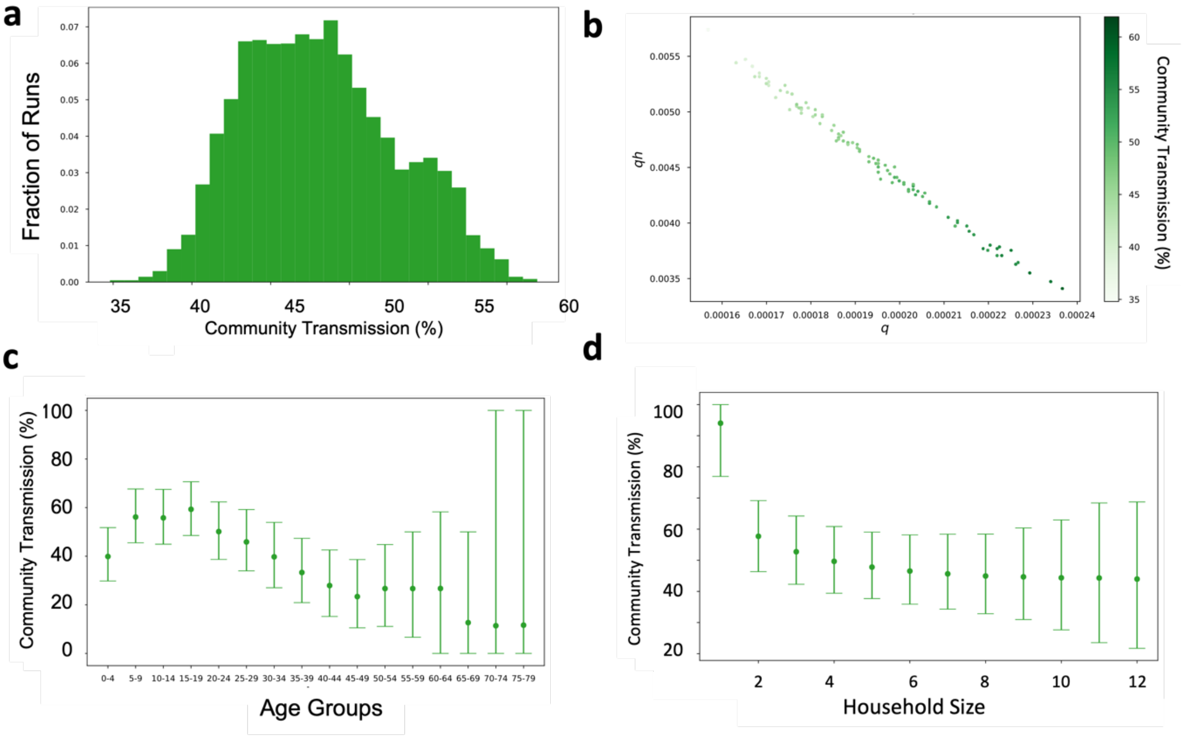

*Figure S10: (a) Histogram of overall community transmission percentage, (b) distribution of overall community transmission percentage, (c) mean and 2.5–97.5% quantiles of community transmission percentages in age groups and (d) mean and 2.5–97.5% quantiles of community transmission percentages in household size groups* ***with an average of 150 days of infestation duration****. Data is captured quarterly after year 5 in 100 simulations.*

Even though decreasing the transmission parameters proportionate to the increase in the duration of infestation was necessary to keep the endemic prevalence or $R_0$ at the same level, the decrease in the transmission parameters makes a difference when it comes to interventions (Figure S11). When the transmission coefficients are lower (higher duration of infestation), the disease spreads more slowly in the community. Therefore, repetitive MDA rounds became more effective under the assumption of lower transmission coefficients (higher duration of infestation) (Figure S12, Figure S13, Table S3, and Table S4).

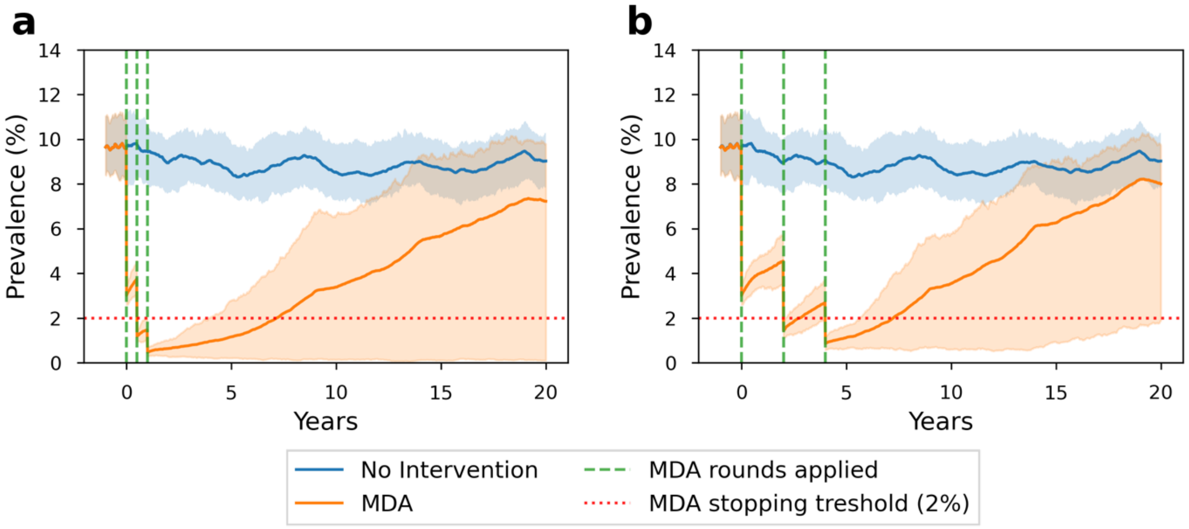

*Figure S11: The scabies prevalence in 20 years with MDA strategies consisting of 3 rounds, 80% population coverage, random individual selection, and (a) six-month (b) two-year time intervals* ***with an average of duration of infestation of 150***

***days***. *The green dashed lines show when MDA rounds are applied. The red dotted lines represent MDA stopping threshold (2%). It is assumed that there is no scabies importation.*

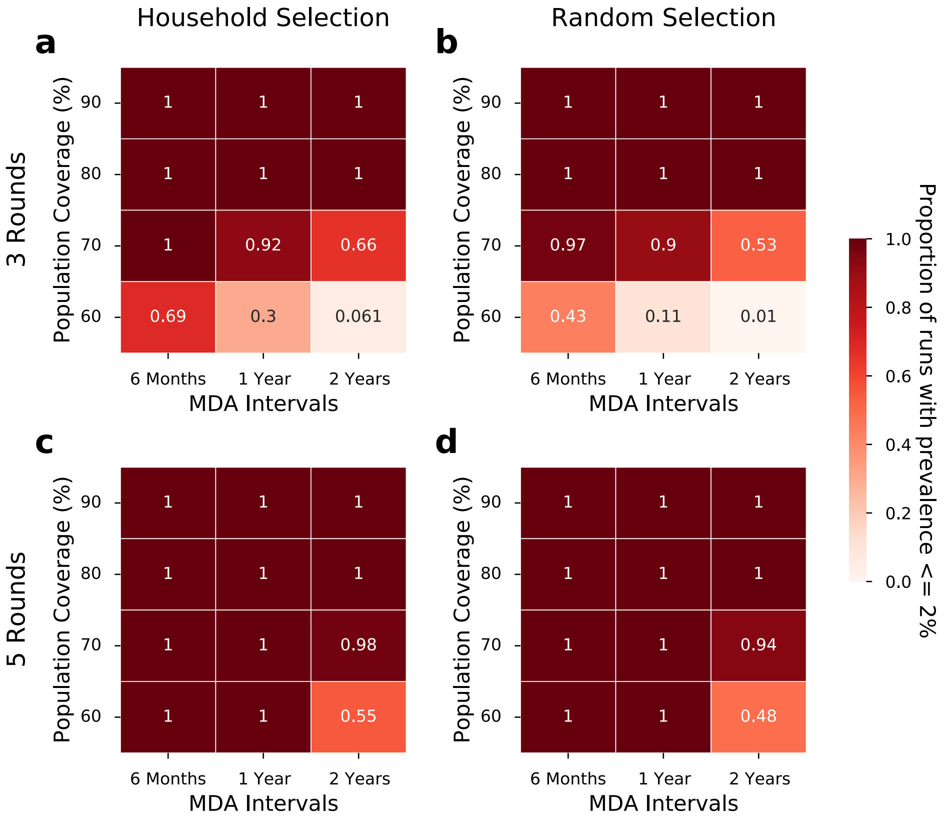

*Figure S12: The proportion of simulation runs with less than 2% prevalence is achieved in differing MDA strategies* ***with an average of duration of infestation of 150 days****. The first column (a & c) are the MDA strategies with household-based selection and the second column (b & d) are the MDA strategies with random individual selection. The first row (a & b) are the MDA strategies with three rounds and the second row (c & d) are the MDA strategies with 5 rounds. Each panel is grouped by the population coverage in MDAs and MDA intervals. Each value is calculated from 100 simulation runs. In these scenarios, it is assumed that there is no scabies importation.*

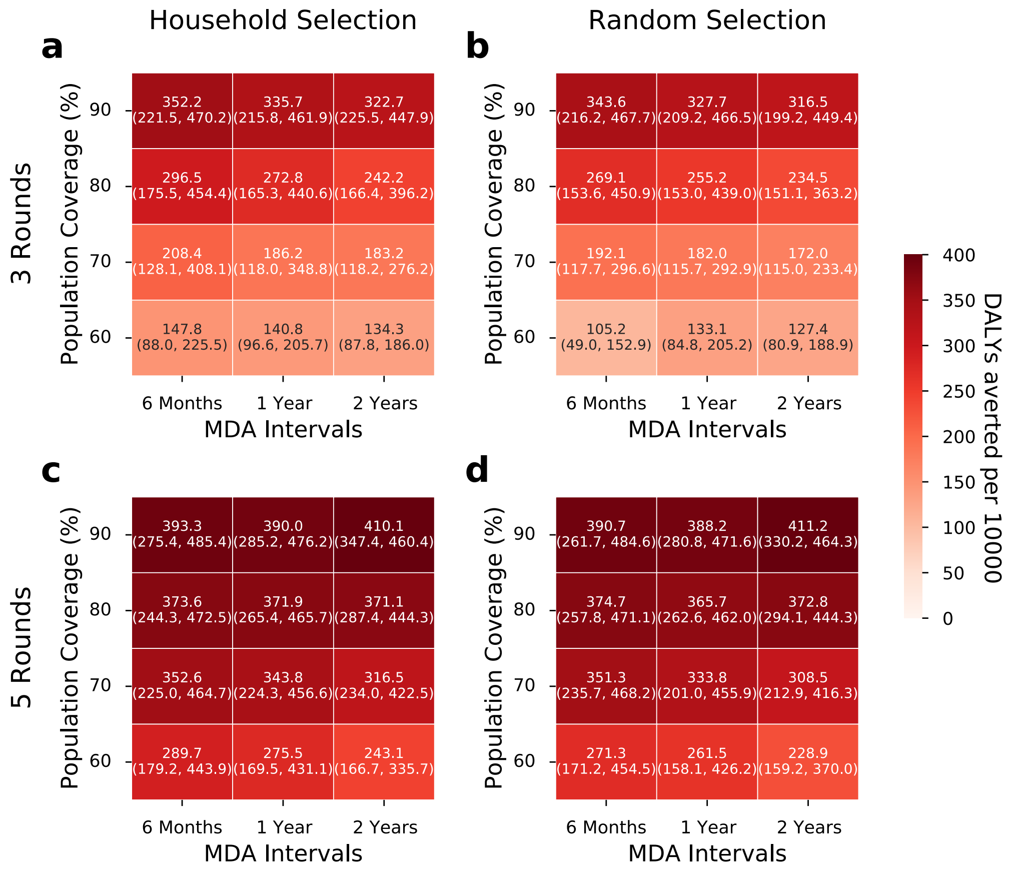

*Figure S13: Mean and 2.5–97.5 quantiles of DALYs averted per 10000 people in differing MDA strategies **with an average duration of infestation of 150 days**. The values are calculated in 20 years starting from the first MDA round. The first column (a & c) are the MDA strategies with household-based selection and the second column (b & d) are the MDA strategies with random individual selection. The first row (a & b) are the MDA strategies with three rounds and the second row (c & d) are the MDA strategies with 5 rounds. Each panel is grouped by the population coverage in MDAs and MDA intervals. Each value is calculated from 100 simulation runs. In these scenarios, it is assumed that there is no scabies importation.*

*Table S3: Percentage of simulations with <=2% prevalence 1 year after the last MDA round, percentage of simulations with scabies elimination, DALYs averted per 10,000 people (mean, 2.5–97.5 quantiles), and time until prevalence returns to baseline are presented in **MDA strategies with 3 rounds with an average duration of infestation of 150 days**. Time until prevalence returns to baseline is calculated among the runs in which scabies is not eliminated. It is assumed that there is no scabies importation. MDA strategies are ordered from best to worst in terms of DALYs averted.*

| Population selection | Population coverage (%) | MDA Interval | The percentage of runs with <=2% prevalence 1 year after the last MDA round (%) | Percentage of runs with scabies elimination (%) | DALYs averted per 10,000 people in 20 years (Mean, 2.5 – 97.5 quantiles) | Time until prevalence returns to baseline (years) |
|---|---|---|---|---|---|---|
| Household | 90 | 6 months | 100 | 0 | 352.2 (221.5, 470.2) | 20+ (15.9, 20+) |
| Random | 90 | 6 months | 100 | 0 | 343.6 (216.2, 467.7) | 20+ (16.0, 20+) |
| Household | 90 | 1 year | 100 | 0 | 335.7 (215.8, 461.9) | 20+ (15.4, 20+) |
| Random | 90 | 1 year | 100 | 0 | 327.7 (209.2, 466.5) | 20+ (14.1, 20+) |
| Household | 90 | 2 years | 100 | 0 | 322.7 (225.5, 447.9) | 20+ (15.6, 20+) |
| Random | 90 | 2 years | 100 | 0 | 316.5 (199.2, 449.4) | 20+ (15.4, 20+) |
| Household | 80 | 6 months | 100 | 0 | 296.5 (175.5, 454.4) | 20+ (13.0, 20+) |
| Household | 80 | 1 year | 100 | 0 | 272.8 (165.3, 440.6) | 20+ (13.0, 20+) |
| Random | 80 | 6 months | 100 | 0 | 269.1 (153.6, 450.9) | 20+ (12.7, 20+) |

| | | | | | | |
|---|---|---|---|---|---|---|
| Random | 80 | 1 year | 100 | 0 | 255.2 (153.0, 439.0) | 20+ (12.7, 20+) |
| Household | 80 | 2 years | 100 | 0 | 242.2 (166.4, 396.2) | 18.9 (13.1, 20+) |
| Random | 80 | 2 years | 100 | 0 | 234.5 (151.1, 363.2) | 18.7 (13.2, 20+) |
| Household | 70 | 6 months | 100 | 0 | 208.4 (128.1, 408.1) | 17.9 (11.3, 20+) |
| Random | 70 | 6 months | 97 | 0 | 192.1 (117.7, 296.6) | 17.0 (11.1, 20+) |
| Household | 70 | 1 year | 92 | 0 | 186.2 (118.0, 348.8) | 17.1 (11.4, 20+) |
| Household | 70 | 2 years | 66 | 0 | 183.2 (118.2, 276.2) | 17.2 (11.4, 20+) |
| Random | 70 | 1 year | 90 | 0 | 182 (115.7, 292.9) | 16.4 (12.4, 20+) |
| Random | 70 | 2 years | 53 | 0 | 172 (115.0, 233.4) | 16.3 (12.3, 20+) |
| Household | 60 | 6 months | 69 | 0 | 147.8 (88.0, 225.5) | 13.6 (8.3, 20+) |
| Household | 60 | 1 year | 30 | 0 | 140.8 (96.6, 205.7) | 13.4 (8.3, 20+) |
| Household | 60 | 2 years | 6 | 0 | 134.3 (87.8, 186.0) | 13.4 (9.3, 20+) |
| Random | 60 | 1 year | 11 | 0 | 133.1 (84.8, 205.2) | 13.3 (8.2, 20+) |
| Random | 60 | 2 years | 1 | 0 | 127.4 (80.9, 188.9) | 13.3 (8.7, 20+) |
| Random | 60 | 6 months | 43 | 0 | 105.2 (49.0, 152.9) | 9.7 (6.7, 20+) |

*Table S4: Percentage of simulations with <=2% prevalence 1 year after the last MDA round, percentage of simulations with scabies elimination, DALYs averted per 10,000 people (mean, 2.5–97.5 quantiles), and time until prevalence returns to baseline are presented in* ***MDA strategies with 5 rounds with an average of duration of infestation of 150 days.*** *Time until prevalence returns to baseline is calculated among the runs in which scabies is not eliminated. It is assumed that there is no scabies importation. MDA strategies are ordered from best to worst in terms of DALYs averted.*

| **Population selection** | **Population coverage (%)** | **MDA Interval** | **The percentage of runs with <=2% prevalence 1 year after the last MDA round (%)** | **Percentag e of runs with scabies eliminatio n (%)** | **DALYs averted per 10,000 people in 20 years (Mean, 2.5 – 97.5 quantiles)** | **Time until prevalence returns to baseline (years)** |
|---|---|---|---|---|---|---|
| Random | 90 | 2 years | 100 | 1 | 411.2 (330.2, 464.3) | 20+ (20.0, 20+) |
| Household | 90 | 2 years | 100 | 2 | 410.1 (347.4, 460.4) | 20+ (20.0, 20+) |
| Household | 90 | 6 months | 100 | 12 | 393.3 (275.4, 485.4) | 20+ (17.9, 20+) |
| Random | 90 | 6 months | 100 | 9 | 390.7 (261.7, 484.6) | 20+ (16.8, 20+) |
| Household | 90 | 1 year | 100 | 9 | 390 (285.2, 476.2) | 20+ (17.8, 20+) |
| Random | 90 | 1 year | 100 | 3 | 388.2 (280.8, 471.6) | 20+ (17.7, 20+) |
| Random | 80 | 6 months | 100 | 0 | 374.7 (257.8, 471.1) | 20+ (17.5, 20+) |
| Household | 80 | 6 months | 100 | 1 | 373.6 (244.3, 472.5) | 20+ (16.0, 20+) |
| Random | 80 | 2 years | 100 | 1 | 372.8 (294.1, 444.3) | 20+ (18.7, 20+) |
| Household | 80 | 1 year | 100 | 1 | 371.9 (265.4, 465.7) | 20+ (17.8, 20+) |
| Household | 80 | 2 years | 100 | 0 | 371.1 (287.4, 444.3) | 20+ (19.8, 20+) |
| Random | 80 | 1 year | 100 | 2 | 365.7 (262.6, 462.0) | 20+ (16.4, 20+) |
| Household | 70 | 6 months | 100 | 0 | 352.6 (225.0, 464.7) | 20+ (14.0, 20+) |
| Random | 70 | 6 months | 100 | 1 | 351.3 (235.7, 468.2) | 20+ (16.6, 20+) |
| Household | 70 | 1 year | 100 | 1 | 343.8 (224.3, 456.6) | 20+ (16.6, 20+) |
| Random | 70 | 1 year | 100 | 0 | 333.8 (201.0, 455.9) | 20+ (14.8, 20+) |
| Household | 70 | 2 years | 98 | 0 | 316.5 (234.0, 422.5) | 20+ (16.7, 20+) |
| Random | 70 | 2 years | 94 | 0 | 308.5 (212.9, 416.3) | 20+ (17.0, 20+) |
| Household | 60 | 6 months | 100 | 0 | 289.7 (179.2, 443.9) | 20+ (13.3, 20+) |
| Household | 60 | 1 year | 100 | 0 | 275.5 (169.5, 431.1) | 20+ (13.7, 20+) |
| Random | 60 | 6 months | 100 | 0 | 271.3 (171.2, 454.5) | 20+ (12.8, 20+) |
| Random | 60 | 1 year | 100 | 0 | 261.5 (158.1, 426.2) | 20+ (13.2, 20+) |
| Household | 60 | 2 years | 55 | 0 | 243.1 (166.7, 335.7) | 20+ (14.8, 20+) |

| | | | | | | |
|---|---|---|---|---|---|---|
| Random | 60 | 2 years | 48 | 0 | 228.9 (159.2, 370.0) | 20+ (15.2, 20+) |

### S6. Additional MDA results

In this section, we present DALYs averted heatmap plot (Figure S14) and tables containing the results of all the MDA scenarios including the ones with 60% and 70% population coverage (Table S5 and Table S6). In section S6.1, we provide results with treatment efficacies of 85% (Section S6.1.1) and 95% (Section S.1.2).

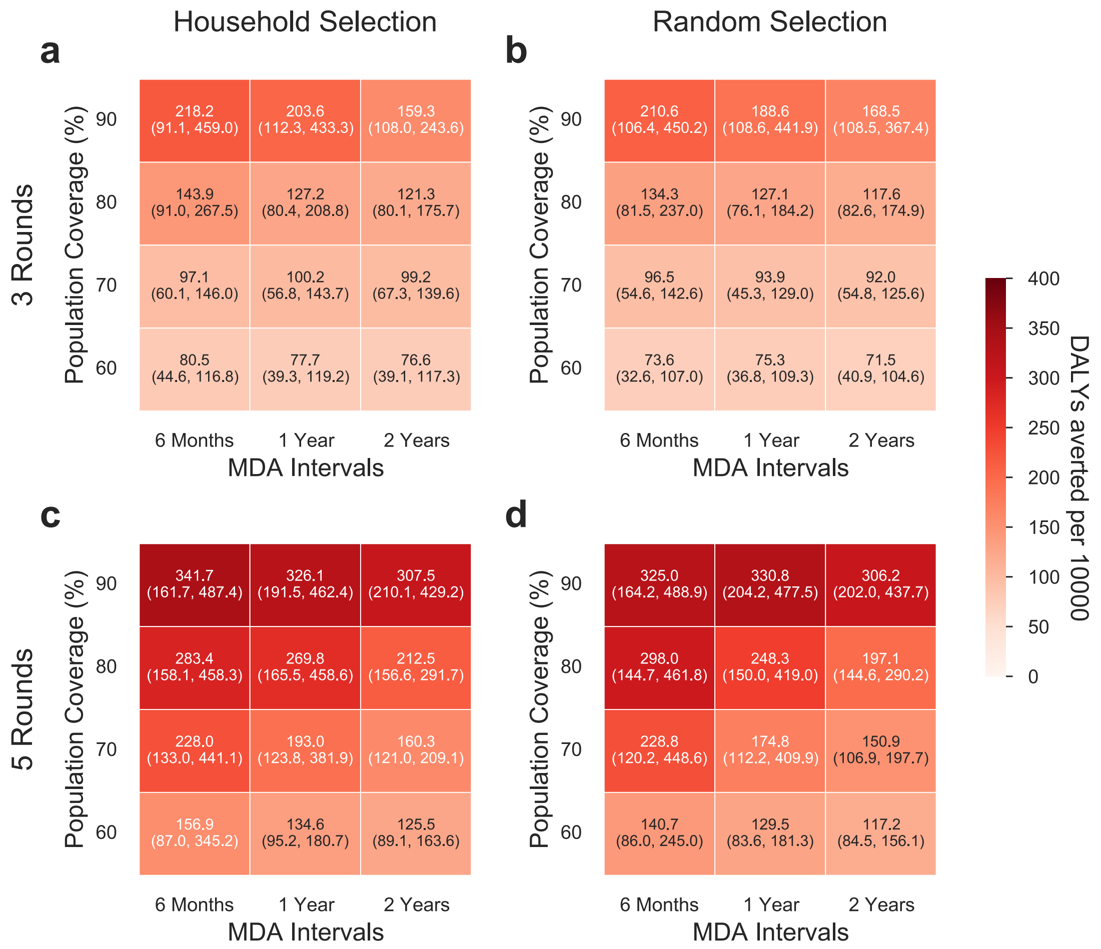

*Figure S14: Mean and 2.5–97.5 quantiles of DALYs averted per 10000 people in differing MDA strategies with* ***an average duration of infestation of 90 days****. The values are calculated in 20 years starting from the first MDA round. The first column (a & c) are the MDA strategies with household-based selection and the second column (b & d) are the MDA strategies with random individual selection. The first row (a & b) are the MDA strategies with three rounds and the second row (c & d) are the MDA strategies with 5 rounds. Each panel is grouped by the population coverage in MDAs and MDA intervals. Each value is calculated from 100 simulation runs. In these scenarios, it is assumed that there is no scabies importation.*

*Table S5: Percentage of simulations with <=2% prevalence 1 year after the last MDA round, percentage of simulations with scabies elimination, DALYs averted per 10,000 people (mean, 2.5–97.5 quantiles), and time until prevalence returns to baseline are presented in **MDA strategies with 3 rounds in scenarios with an average duration of infestation of 90 days**. Time until prevalence returns to baseline is calculated among the runs in which scabies is not eliminated. It is assumed that there is no scabies importation. MDA strategies are ordered from best to worst in terms of DALYs averted.*

| Population selection | Population coverage (%) | MDA Interval | The percentage of runs with <=2% prevalence 1 year after the last MDA round (%) | Percentage of runs with scabies elimination (%) | DALYs averted per 10,000 people in 20 years (Mean, 2.5 – 97.5 quantiles) | Time until prevalence returns to baseline (years) |
|---|---|---|---|---|---|---|
| Household | 90 | 6 months | 100 | 0 | 218.2 (91.1, 459.0) | 18.4 (8.2, 20+) |
| Random | 90 | 6 months | 100 | 0 | 210.6 (106.4, 450.2) | 17.7 (7.7, 20+) |
| Household | 90 | 1 year | 100 | 0 | 203.6 (112.3, 433.3) | 17.7 (8.1, 20+) |
| Random | 90 | 1 year | 100 | 0 | 188.6 (108.6, 441.9) | 13.6 (8.1, 20+) |
| Random | 90 | 2 years | 91 | 0 | 168.5 (108.5, 367.4) | 13.2 (8.5, 20+) |
| Household | 90 | 2 years | 92 | 0 | 159.3 (108.0, 243.6) | 12.8 (8.7, 17.8) |
| Household | 80 | 6 months | 99 | 0 | 143.9 (91.0, 267.5) | 12.3 (6.9, 17.9) |
| Random | 80 | 6 months | 100 | 0 | 134.3 (81.5, 237.0) | 12.2 (6.8, 18.4) |
| Household | 80 | 1 year | 78 | 0 | 127.2 (80.4, 208.8) | 11.8 (7.1, 13.8) |
| Random | 80 | 1 year | 76 | 0 | 127.1 (76.1, 184.2) | 11.2 (7.2, 20+) |
| Household | 80 | 2 years | 35 | 0 | 121.3 (80.1, 175.7) | 11.4 (8.0, 19.0) |
| Random | 80 | 2 years | 26 | 0 | 117.6 (82.6, 174.9) | 11.4 (7.6, 17.9) |
| Household | 70 | 1 year | 21 | 0 | 100.2 (56.8, 143.7) | 8.2 (6.4, 13.4) |
| Household | 70 | 2 years | 2 | 0 | 99.2 (67.3, 139.6) | 8.8 (7.6, 13.5) |
| Household | 70 | 6 months | 69 | 0 | 97.1 (60.1, 146.0) | 8.0 (6.2, 13.3) |
| Random | 70 | 6 months | 58 | 0 | 96.5 (54.6, 142.6) | 8.0 (6.2, 13.2) |
| Random | 70 | 1 year | 12 | 0 | 93.9 (45.3, 129.0) | 8.0 (6.5, 13.9) |
| Random | 70 | 2 years | 0 | 0 | 92 (54.8, 125.6) | 8.6 (7.3, 13.2) |
| Household | 60 | 6 months | 16 | 0 | 80.5 (44.6, 116.8) | 7.5 (5.6, 13.5) |
| Household | 60 | 1 year | 0 | 0 | 77.7 (39.3, 119.2) | 7.5 (5.7, 13.7) |
| Household | 60 | 2 years | 0 | 0 | 76.6 (39.1, 117.3) | 8.1 (1.5, 13.8) |
| Random | 60 | 1 year | 0 | 0 | 75.3 (36.8, 109.3) | 7.3 (5.6, 13.4) |
| Random | 60 | 6 months | 4 | 0 | 73.6 (32.6, 107.0) | 7.1 (5.7, 13.0) |
| Random | 60 | 2 years | 0 | 0 | 71.5 (40.9, 104.6) | 7.9 (1.5, 13.3) |

*Table S6: Percentage of simulations with <=2% prevalence 1 year after the last MDA round, percentage of simulations with scabies elimination, DALYs averted per 10,000 people (mean, 2.5–97.5 quantiles), and time until prevalence returns to baseline are presented in* ***MDA strategies with 5 rounds in scenarios with an average duration of infestation of 90 days****. Time until prevalence returns to baseline is calculated among the runs in which scabies is not eliminated. It is assumed that there is no scabies importation. MDA strategies are ordered from best to worst in terms of DALYs averted.*

| Population selection | Population coverage (%) | MDA Interval | The percentage of runs with <=2% prevalence 1 year after the last MDA round (%) | Percentage of runs with scabies elimination (%) | DALYs averted per 10,000 people in 20 years (Mean, 2.5 – 97.5 quantiles) | Time until prevalence returns to baseline (years) |
|---|---|---|---|---|---|---|
| Household | 90 | 6 months | 100 | 18 | 341.7 (161.7, 487.4) | 20+ (11.6, 20+) |
| Random | 90 | 1 year | 100 | 10 | 330.8 (204.2, 477.5) | 20+ (12.7, 20+) |
| Household | 90 | 1 year | 100 | 10 | 326.1 (191.5, 462.4) | 20+ (12.7, 20+) |
| Random | 90 | 6 months | 100 | 16 | 325 (164.2, 488.9) | 20+ (11.7, 20+) |
| Household | 90 | 2 years | 99 | 0 | 307.5 (210.1, 429.2) | 20+ (13.8, 20+) |
| Random | 90 | 2 years | 98 | 3 | 306.2 (202.0, 437.7) | 20+ (13.5, 20+) |
| Random | 80 | 6 months | 100 | 0 | 298 (144.7, 461.8) | 20+ (11.2, 20+) |
| Household | 80 | 6 months | 100 | 0 | 283.4 (158.1, 458.3) | 20+ (11.6, 20+) |
| Household | 80 | 1 year | 100 | 0 | 269.8 (165.5, 458.6) | 20+ (11.5, 20+) |
| Random | 80 | 1 year | 100 | 0 | 248.3 (150.0, 419.0) | 18.2 (11.7, 20+) |
| Random | 70 | 6 months | 100 | 0 | 228.8 (120.2, 448.6) | 18.0 (8.7, 20+) |
| Household | 70 | 6 months | 100 | 0 | 228 (133.0, 441.1) | 18.2 (8.5, 20+) |
| Household | 80 | 2 years | 64 | 0 | 212.5 (156.6, 291.7) | 16.4 (12.9, 20+) |
| Random | 80 | 2 years | 46 | 0 | 197.1 (144.6, 290.2) | 15.9 (12.4, 20+) |
| Household | 70 | 1 year | 97 | 0 | 193 (123.8, 381.9) | 13.7 (10.8, 20+) |
| Random | 70 | 1 year | 98 | 0 | 174.8 (112.2, 409.9) | 13.2 (8.6, 20+) |
| Household | 70 | 2 years | 6 | 0 | 160.3 (121.0, 209.1) | 13.5 (11.8, 18.3) |
| Household | 60 | 6 months | 98 | 0 | 156.9 (87.0, 345.2) | 12.9 (7.8, 20+) |
| Random | 70 | 2 years | 3 | 0 | 150.9 (106.9, 197.7) | 13.3 (11.9, 17.6) |
| Random | 60 | 6 months | 91 | 0 | 140.7 (86.0, 245.0) | 12.3 (7.2, 18.9) |
| Household | 60 | 1 year | 67 | 0 | 134.6 (95.2, 180.7) | 12.2 (8.4, 17.7) |
| Random | 60 | 1 year | 38 | 0 | 129.5 (83.6, 181.3) | 12.1 (8.0, 18.4) |
| Household | 60 | 2 years | 0 | 0 | 125.5 (89.1, 163.6) | 12.9 (1.5, 17.7) |
| Random | 60 | 2 years | 0 | 0 | 117.2 (84.5, 156.1) | 12.8 (1.5, 18.7) |

#### S6.1. Results with various treatment efficacies

In the main manuscript, we present results of MDA strategies with 90% treatment efficacy. In this section, we present the comparison of MDA strategies under the assumption of 85% treatment efficacy (Section S6.1.1) and 95% treatment efficacy (Section S6.1.2).

##### S6.1.1. Results with treatment efficacy of 85% (rather than 90%)

In MDA strategies with treatment efficacy of 85% (rather than 90%), 80% population coverage, household selection in 5 rounds with 2-year time interval, the probability of achieving a prevalence of less than the stopping threshold decreases from 0.64 to 0.24 (Figure S15). Average DALYs averted per 10,000 people in 20 years decreases from 80.5 years to 75 years in MDA strategies with 60% coverage, household-based individual selection in 3 rounds with 6-month time interval when a treatment efficacy of 85% is assumed rather than 90% (Table S7). The probability of scabies elimination decreases from 18% to 5% in MDA strategies with 90% coverage, household-based individual selection in 5 rounds with 6-month time interval when a treatment efficacy of 85% is assumed rather than 90% (Table S8).

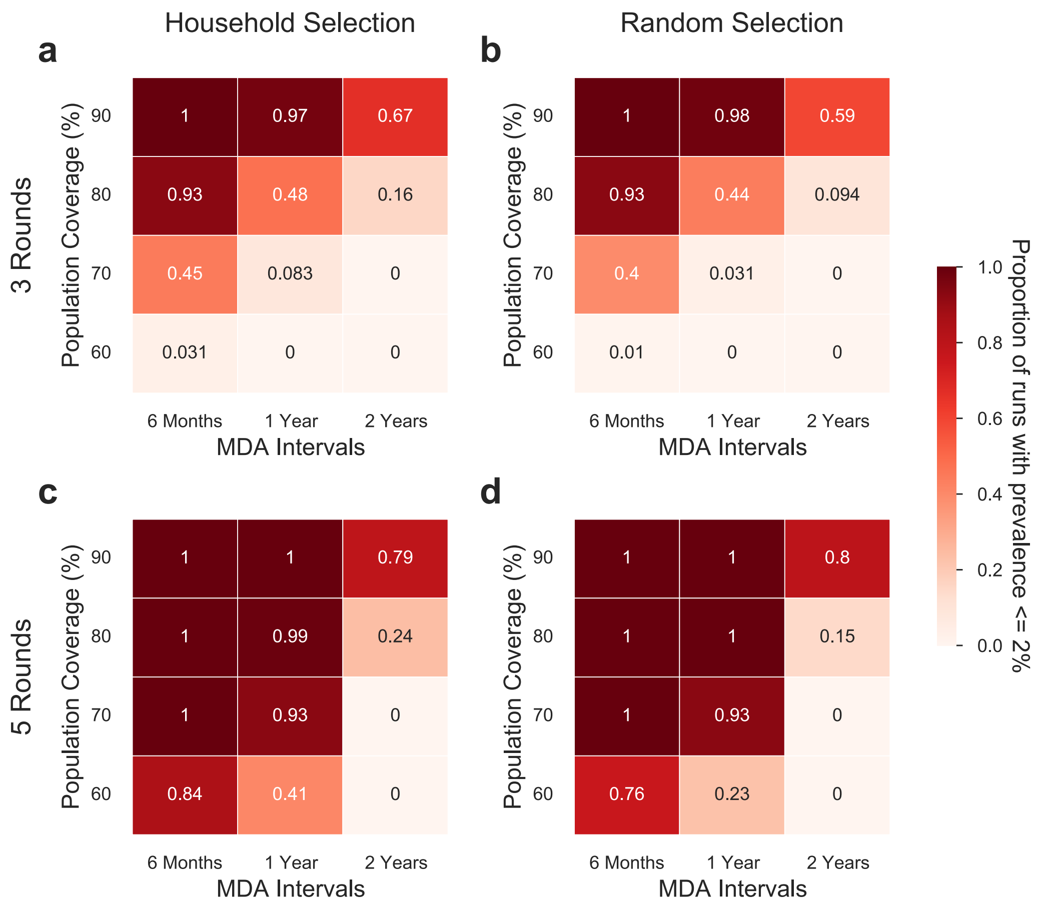

*Figure S15: The proportion of simulation runs with a prevalence of less than 2% is achieved in differing MDA strategies having a treatment efficacy of 85% (rather than 90%) in scenarios with **an average duration of infestation of 90 days**. The first column (a & c) are the MDA strategies with household-based selection and the second column (b & d) are the MDA strategies with random individual selection. The first row (a & b) are the MDA strategies with three rounds and the second row (c & d) are the MDA strategies with 5 rounds. Each panel is grouped by the population coverage in MDAs and MDA intervals. Each value is calculated from 100 simulation runs. It is assumed that there is no scabies importation.*

*Table S7: Percentage of simulations with <=2% prevalence 1 year after the last MDA round, percentage of simulations with scabies elimination, DALYs averted per 10,000 people (mean, 2.5–97.5 quantiles), and time until prevalence returns to baseline are presented in **MDA strategies with 3 rounds and 85% treatment efficacy** in scenarios with **an average duration*** *of infestation of 90 days. Time until prevalence returns to baseline is calculated among the runs in which scabies is not eliminated. It is assumed that there is no scabies importation. MDA strategies are ordered from best to worst in terms of DALYs averted.*

| Population selection | Population coverage (%) | MDA Interval | The percentage of runs with <=2% prevalence 1 year after the last MDA round (%) | Percentage of runs with scabies elimination (%) | DALYs averted per 10,000 people in 20 years (Mean, 2.5–97.5 quantiles) | Time until prevalence returns to baseline (years) |
|---|---|---|---|---|---|---|
| Household | 90 | 6 months | 100 | 0 | 182.8 (89.1, 435.4) | 13.6 (7.6, 20.0) |
| Random | 90 | 6 months | 100 | 0 | 161.5 (92.9, 393.4) | 12.9 (7.2, 20.0) |
| Random | 90 | 1 year | 98 | 0 | 145.5 (92.3, 253.4) | 12.5 (7.6, 18.1) |
| Household | 90 | 1 year | 97 | 0 | 144 (97.2, 214.1) | 12.2 (7.7, 18.1) |
| Household | 90 | 2 years | 67 | 0 | 137.6 (96.5, 189.5) | 12.2 (8.1, 13.8) |
| Random | 90 | 2 years | 59 | 0 | 135.5 (92.0, 182.1) | 12.2 (7.8, 13.9) |
| Household | 80 | 6 months | 93 | 0 | 118.4 (79.7, 173.1) | 8.8 (6.9, 17.1) |
| Random | 80 | 6 months | 93 | 0 | 115 (64.5, 167.6) | 8.7 (6.6, 18.5) |
| Household | 80 | 1 year | 48 | 0 | 110 (69.7, 152.9) | 8.6 (6.6, 13.4) |
| Household | 80 | 2 years | 16 | 0 | 109.7 (75.5, 155.3) | 11.2 (7.8, 13.7) |
| Random | 80 | 1 year | 44 | 0 | 106.1 (70.9, 143.2) | 8.4 (6.6, 13.5) |
| Random | 80 | 2 years | 9 | 0 | 104.5 (68.2, 148.9) | 11.1 (7.5, 13.5) |
| Household | 70 | 6 months | 45 | 0 | 92.5 (50.2, 141.8) | 7.8 (6.2, 13.8) |
| Random | 70 | 6 months | 40 | 0 | 89.3 (53.0, 138.1) | 7.7 (5.9, 13.4) |
| Household | 70 | 2 years | 0 | 0 | 87.9 (54.6, 132.0) | 8.5 (1.7, 13.5) |
| Household | 70 | 1 year | 8 | 0 | 87.3 (50.6, 124.9) | 7.8 (6.1, 13.3) |
| Random | 70 | 1 year | 3 | 0 | 87.3 (49.7, 122.6) | 7.7 (6.4, 13.7) |
| Random | 70 | 2 years | 0 | 0 | 84 (53.1, 124.5) | 8.5 (1.6, 13.6) |
| Household | 60 | 6 months | 3 | 0 | 75 (40.9, 112.9) | 7.0 (5.1, 13.8) |
| Household | 60 | 1 year | 0 | 0 | 74.7 (34.8, 111.0) | 7.3 (5.8, 13.4) |
| Household | 60 | 2 years | 0 | 0 | 71.6 (36.9, 105.3) | 7.8 (1.5, 13.8) |
| Random | 60 | 1 year | 0 | 0 | 70.2 (34.3, 104.0) | 7.0 (5.4, 13.5) |
| Random | 60 | 6 months | 1 | 0 | 67.8 (35.2, 105.2) | 6.9 (4.4, 13.3) |
| Random | 60 | 2 years | 0 | 0 | 66.8 (32.9, 106.2) | 7.8 (1.5, 13.3) |

*Table S8: Percentage of simulations with <=2% prevalence 1 year after the last MDA round, percentage of simulations with scabies elimination, DALYs averted per 10,000 people (mean, 2.5 – 97.5 quantiles), and time until prevalence returns to baseline are presented in **MDA strategies with 5 rounds and 85% treatment efficacy** in scenarios with **an average duration of infestation of 90 days.** Time until prevalence returns to baseline is calculated among the runs in which scabies is not eliminated. It is assumed that there is no scabies importation. MDA strategies are ordered from best to worst in terms of DALYs averted.*

| Population selection | Population coverage (%) | MDA Interval | The percentage of runs with <=2% prevalence 1 year after the last MDA round (%) | Percentage of runs with scabies elimination (%) | DALYs averted per 10,000 people in 20 years (Mean, 2.5 – 97.5 quantiles) | Time until prevalence returns to baseline (years) |
|---|---|---|---|---|---|---|
| Household | 90 | 6 months | 100 | 5 | 319.3 (157.4, 480.5) | 20+ (11.4, 20+) |
| Random | 90 | 6 months | 100 | 11 | 313.9 (159.4, 484.8) | 20+ (9.0, 20+) |
| Household | 90 | 1 year | 100 | 3 | 295.3 (184.9, 461.3) | 20+ (11.8, 20+) |

| | | | | | | |
|---|---|---|---|---|---|---|
| Random | 90 | 1 year | 100 | 5 | 290.6 (175.0, 467.1) | 20+ (12.3, 20+) |
| Household | 80 | 6 months | 100 | 0 | 275.6 (144.8, 477.6) | 20+ (8.9, 20+) |
| Random | 80 | 6 months | 100 | 0 | 263.7 (115.8, 462.4) | 20+ (8.5, 20+) |
| Household | 90 | 2 years | 79 | 0 | 246.2 (178.6, 405.3) | 18.1 (13.3, 20+) |
| Random | 90 | 2 years | 80 | 0 | 241.3 (167.8, 384.6) | 17.9 (12.7, 20+) |
| Random | 80 | 1 year | 100 | 0 | 235.8 (141.7, 428.9) | 18.2 (11.1, 20+) |
| Household | 80 | 1 year | 99 | 0 | 225 (130.7, 413.9) | 17.7 (11.0, 20+) |
| Household | 70 | 6 months | 100 | 0 | 207 (112.0, 421.9) | 17.2 (8.1, 20+) |
| Random | 70 | 6 months | 100 | 0 | 193.1 (113.2, 420.3) | 13.9 (8.1, 20+) |
| Household | 80 | 2 years | 24 | 0 | 183 (138.3, 251.8) | 13.9 (12.2, 18.6) |
| Random | 80 | 2 years | 15 | 0 | 177 (134.4, 236.4) | 13.8 (12.2, 20+) |
| Random | 70 | 1 year | 93 | 0 | 162.5 (107.1, 249.3) | 12.9 (8.7, 20+) |
| Household | 70 | 1 year | 93 | 0 | 159.4 (113.3, 227.8) | 12.7 (8.7, 18.3) |
| Household | 70 | 2 years | 0 | 0 | 143 (108.1, 191.0) | 13.2 (1.7, 16.4) |
| Random | 70 | 2 years | 0 | 0 | 138.5 (100.3, 178.6) | 13.0 (1.6, 17.9) |
| Household | 60 | 6 months | 84 | 0 | 133.8 (80.9, 213.6) | 12.1 (7.7, 18.4) |
| Household | 60 | 1 year | 41 | 0 | 122.5 (86.8, 165.8) | 11.4 (8.1, 13.7) |
| Random | 60 | 6 months | 76 | 0 | 120.8 (78.4, 176.3) | 11.2 (7.0, 18.5) |
| Household | 60 | 2 years | 0 | 0 | 117.3 (78.7, 155.9) | 12.8 (1.5, 18.8) |
| Random | 60 | 1 year | 23 | 0 | 114.9 (77.6, 152.6) | 11.2 (7.6, 13.4) |
| Random | 60 | 2 years | 0 | 0 | 110.7 (79.6, 141.1) | 12.8 (1.5, 20+) |

#### S6.1.2. Results with treatment efficacy of 95% (rather than 90%)

In MDA strategies with treatment efficacy of 95% (rather than 90%), 80% population coverage, household selection in 5 rounds with 2-year time interval, the probability of achieving a prevalence of less than the stopping threshold increases from 0.64 to 0.89 (Figure S16). Average DALYs averted per 10,000 people in 20 years increases from 80.5 years to 87 years in MDA strategies with 60% coverage, household-based individual selection in 3 rounds with 6-month time interval when a treatment efficacy of 95% is assumed rather than 90% (Table S9). The probability of scabies elimination increases from 18% to 22% in MDA strategies with 90% coverage, household-based individual selection in 5 rounds with 6-month time interval when a treatment efficacy of 95% is assumed rather than 90% (Table S10).

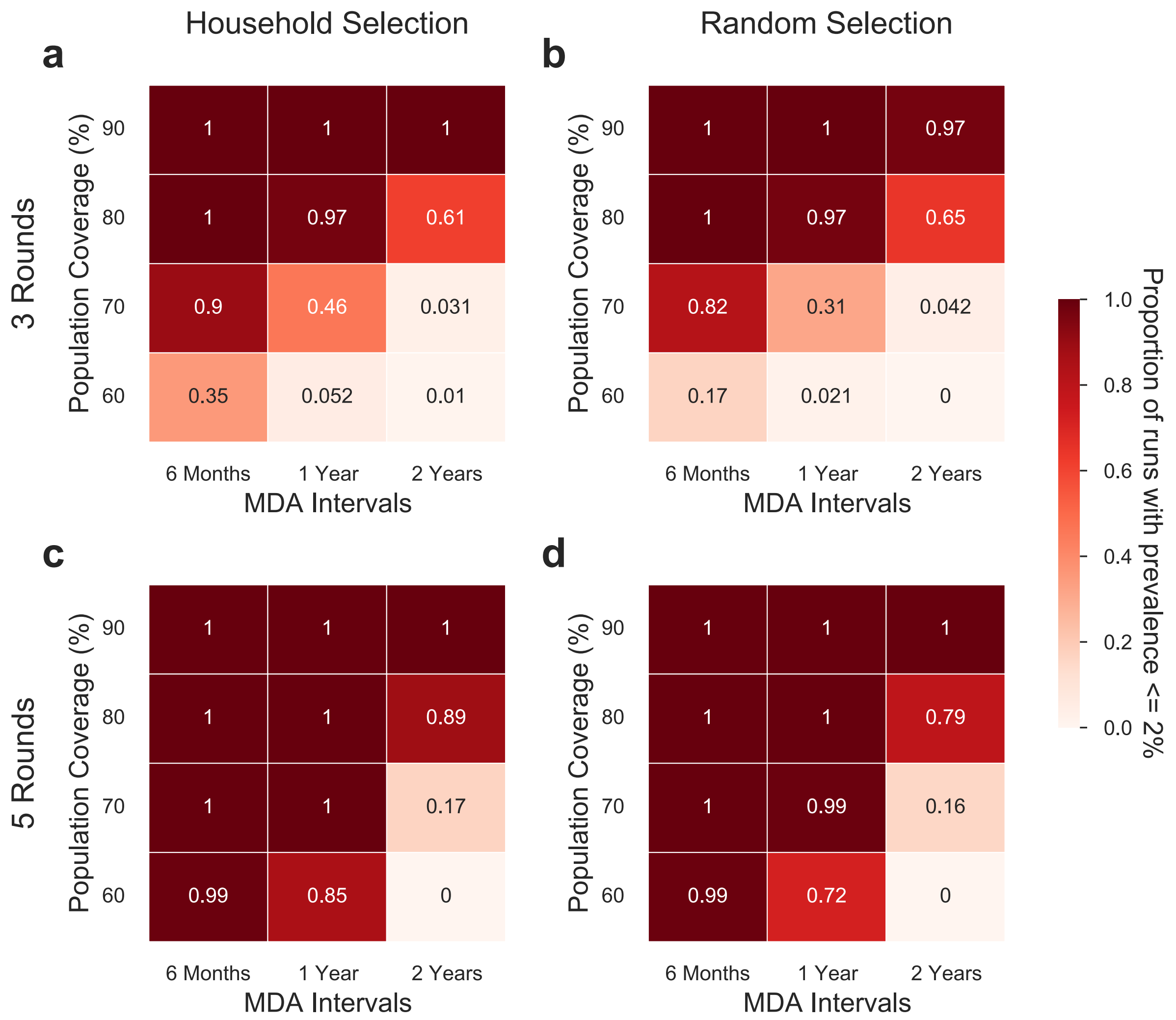

*Figure S16: The proportion of simulation runs with a prevalence of less than or equal to 2% is achieved in differing MDA strategies having a treatment efficacy of 95% (rather than 90%) in scenarios with* ***an average duration of infestation of 90 days****. The first column (a & c) are the MDA strategies with household-based selection and the second column (b & d) are the MDA strategies with random individual selection. The first row (a & b) are the MDA strategies with three rounds and the second row (c & d) are the MDA strategies with 5 rounds. Each panel is grouped by the population coverage in MDAs and MDA intervals. Each value is calculated from 100 simulation runs.*

*Table S9: Percentage of simulations with <=2% prevalence 1 year after the last MDA round, percentage of simulations with scabies elimination, DALYs averted per 10,000 people (mean, 2.5–97.5 quantiles), and time until prevalence returns to baseline are presented in* ***MDA strategies with 3 rounds and 95% treatment efficacy*** *in scenarios with* ***an average duration of infestation of 90 days.*** *Time until prevalence returns to baseline is calculated among the runs in which scabies is not eliminated. It is assumed that there is no scabies importation. MDA strategies are ordered from best to worst in terms of DALYs averted.*

| **Population selection** | **Population coverage (%)** | **MDA Interval** | **The percentage of runs with <=2% prevalence 1 year after the last MDA round (%)** | **Percentage of runs with scabies elimination (%)** | **DALYs averted per 10,000 people in 20 years (Mean, 2.5 – 97.5 quantiles)** | **Time until prevalence returns to baseline (years)** |
|---|---|---|---|---|---|---|
| Household | 90 | 6 months | 100 | 0 | 254.3 (122.7, 481.9) | 20+ (8.6, 20+) |
| Random | 90 | 6 months | 100 | 0 | 253.6 (127.3, 459.7) | 20+ (8.8, 20+) |
| Household | 90 | 1 year | 100 | 0 | 234.7 (120.9, 450.8) | 18.8 (8.9, 20+) |
| Random | 90 | 1 year | 100 | 0 | 228.3 (123.1, 469.6) | 18.0 (8.6, 20+) |

| | | | | | | |
|---|---|---|---|---|---|---|
| Household | 90 | 2 years | 100 | 0 | 199.4 (131.8, 409.3) | 13.9 (10.2, 20+) |
| Random | 90 | 2 years | 97 | 0 | 189.7 (121.7, 381.4) | 13.6 (10.3, 20+) |
| Household | 80 | 6 months | 100 | 0 | 168.5 (90.8, 412.9) | 13.0 (7.6, 20+) |
| Random | 80 | 1 year | 97 | 0 | 156.3 (90.9, 423.5) | 12.9 (7.5, 20+) |
| Household | 80 | 1 year | 97 | 0 | 150.9 (95.2, 284.9) | 12.7 (7.8, 18.8) |
| Random | 80 | 6 months | 100 | 0 | 150.2 (91.1, 226.2) | 12.5 (6.9, 18.0) |
| Household | 80 | 2 years | 61 | 0 | 135.6 (89.5, 189.5) | 12.2 (8.4, 17.5) |
| Random | 80 | 2 years | 65 | 0 | 133.8 (94.0, 181.0) | 12.2 (8.3, 13.7) |
| Household | 70 | 6 months | 90 | 0 | 114.2 (73.5, 183.2) | 8.8 (6.7, 13.5) |
| Random | 70 | 6 months | 82 | 0 | 110.3 (62.2, 162.4) | 8.5 (6.2, 13.6) |
| Household | 70 | 1 year | 46 | 0 | 106.2 (70.5, 146.5) | 8.4 (6.6, 13.6) |
| Household | 70 | 2 years | 3 | 0 | 106 (70.5, 147.7) | 11.0 (7.5, 13.6) |
| Random | 70 | 1 year | 31 | 0 | 102.8 (62.6, 148.9) | 8.3 (6.8, 13.4) |
| Random | 70 | 2 years | 4 | 0 | 102 (68.5, 142.2) | 11.0 (7.6, 13.6) |
| Household | 60 | 6 months | 35 | 0 | 87 (44.5, 120.6) | 7.7 (6.0, 13.7) |
| Household | 60 | 1 year | 5 | 0 | 85.1 (42.5, 134.0) | 7.8 (6.2, 13.6) |
| Household | 60 | 2 years | 1 | 0 | 84.6 (55.3, 119.8) | 8.4 (6.9, 13.5) |
| Random | 60 | 6 months | 17 | 0 | 81.3 (44.7, 117.0) | 7.4 (5.8, 13.1) |
| Random | 60 | 1 year | 2 | 0 | 80.5 (41.6, 121.4) | 7.6 (6.1, 13.8) |
| Random | 60 | 2 years | 0 | 0 | 80.5 (39.8, 113.4) | 8.1 (1.5, 13.2) |

*Table S10: Percentage of simulations with <=2% prevalence 1 year after the last MDA round, percentage of simulations with scabies elimination, DALYs averted per 10,000 people (mean, 2.5 – 97.5 quantiles), and time until prevalence returns to baseline are presented in* ***MDA strategies with 5 rounds and 95% treatment efficacy*** *in scenarios with* ***an average duration of infestation of 90 days.*** *Time until prevalence returns to baseline is calculated among the runs in which scabies is not eliminated. It is assumed that there is no scabies importation. MDA strategies are ordered from best to worst in terms of DALYs averted.*

| Population selection | Population coverage (%) | MDA Interval | The percentage of runs with <=2% prevalence 1 year after the last MDA round (%) | Percentage of runs with scabies elimination (%) | DALYs averted per 10,000 people in 20 years (Mean, 2.5 – 97.5 quantiles) | Time until prevalence returns to baseline (years) |
|---|---|---|---|---|---|---|
| Household | 90 | 2 years | 100 | 6 | 364.5 (235.9, 462.1) | 20+ (15.9, 20+) |
| Household | 90 | 1 year | 100 | 12 | 355.6 (221.8, 481.5) | 20+ (13.5, 20+) |
| Household | 90 | 6 months | 100 | 22 | 345.3 (180.1, 486.8) | 20+ (12.5, 20+) |
| Random | 90 | 1 year | 100 | 16 | 343.3 (196.4, 478.3) | 20+ (12.6, 20+) |
| Random | 90 | 2 years | 100 | 5 | 342.5 (233.4, 458.1) | 20+ (14.8, 20+) |
| Random | 90 | 6 months | 100 | 16 | 342.4 (180.6, 487.2) | 20+ (12.5, 20+) |
| Random | 80 | 6 months | 100 | 0 | 310.2 (152.2, 469.8) | 20+ (11.1, 20+) |
| Household | 80 | 6 months | 100 | 0 | 306 (162.7, 479.8) | 20+ (11.4, 20+) |
| Household | 80 | 1 year | 100 | 0 | 300.5 (172.7, 464.8) | 20+ (12.0, 20+) |
| Random | 80 | 1 year | 100 | 0 | 286 (154.4, 469.0) | 20+ (11.6, 20+) |
| Household | 70 | 6 months | 100 | 0 | 284 (150.3, 478.7) | 20+ (10.6, 20+) |
| Random | 70 | 6 months | 100 | 0 | 262.5 (135.8, 462.8) | 20+ (8.4, 20+) |
| Household | 80 | 2 years | 89 | 0 | 249.8 (174.9, 401.7) | 18.2 (12.8, 20+) |
| Random | 80 | 2 years | 79 | 0 | 238.6 (173.0, 401.6) | 17.8 (12.7, 20+) |
| Household | 70 | 1 year | 100 | 0 | 233.2 (135.9, 460.2) | 18.4 (10.5, 20+) |

| | | | | | | |
|---|---|---|---|---|---|---|
| Random | 70 | 1 year | 99 | 0 | 208.4 (123.5, 405.0) | 16.5 (9.7, 20+) |
| Household | 60 | 6 months | 99 | 0 | 183.5 (98.8, 439.0) | 13.6 (8.1, 20+) |
| Household | 70 | 2 years | 17 | 0 | 176.5 (133.0, 223.0) | 13.8 (12.6, 18.7) |
| Random | 70 | 2 years | 16 | 0 | 168.8 (131.2, 226.7) | 13.6 (11.4, 18.8) |
| Random | 60 | 6 months | 99 | 0 | 163.2 (100.1, 411.6) | 12.9 (7.9, 20+) |
| Household | 60 | 1 year | 85 | 0 | 159.5 (111.3, 250.5) | 13.0 (8.6, 18.6) |
| Household | 60 | 2 years | 0 | 0 | 140.2 (110.3, 180.8) | 13.1 (11.8, 17.7) |
| Random | 60 | 1 year | 72 | 0 | 139.6 (94.9, 197.7) | 12.3 (8.0, 17.8) |
| Random | 60 | 2 years | 0 | 0 | 131.7 (86.7, 182.5) | 13.0 (1.5, 17.7) |

#### S6.2. Results with scabies importation

In this section, we present effectiveness of MDA rounds in scenarios with one person infested by scabies outside of the community every week under the assumption of an average infestation duration of 90 days and 150 days. We compare the results with the ‘no importation’ scenarios (Figure S17 and Figure S18).

We observe that scenarios with and without scabies importation differentiate the most in terms of scabies prevalence at endemic level. When one weekly scabies case is imported to the population through migration, average scabies prevalence increases from 1% to 3% in scenarios with an average infestation duration of 81 days (Figure S17).

While it is possible to expect improvements in the healthcare system in the long run without an MDA strategy, it is unlikely to break the normalisation cycle and expect more people to seek treatment without a capacity problem in the healthcare system. For instance, in order to achieve a 20% reduction in the infestation duration without an MDA (difference between red solid and dotted lines in Figure S17.b), more than 23,000 additional people are needed to seek treatment between year two and three in Monrovia, Liberia.

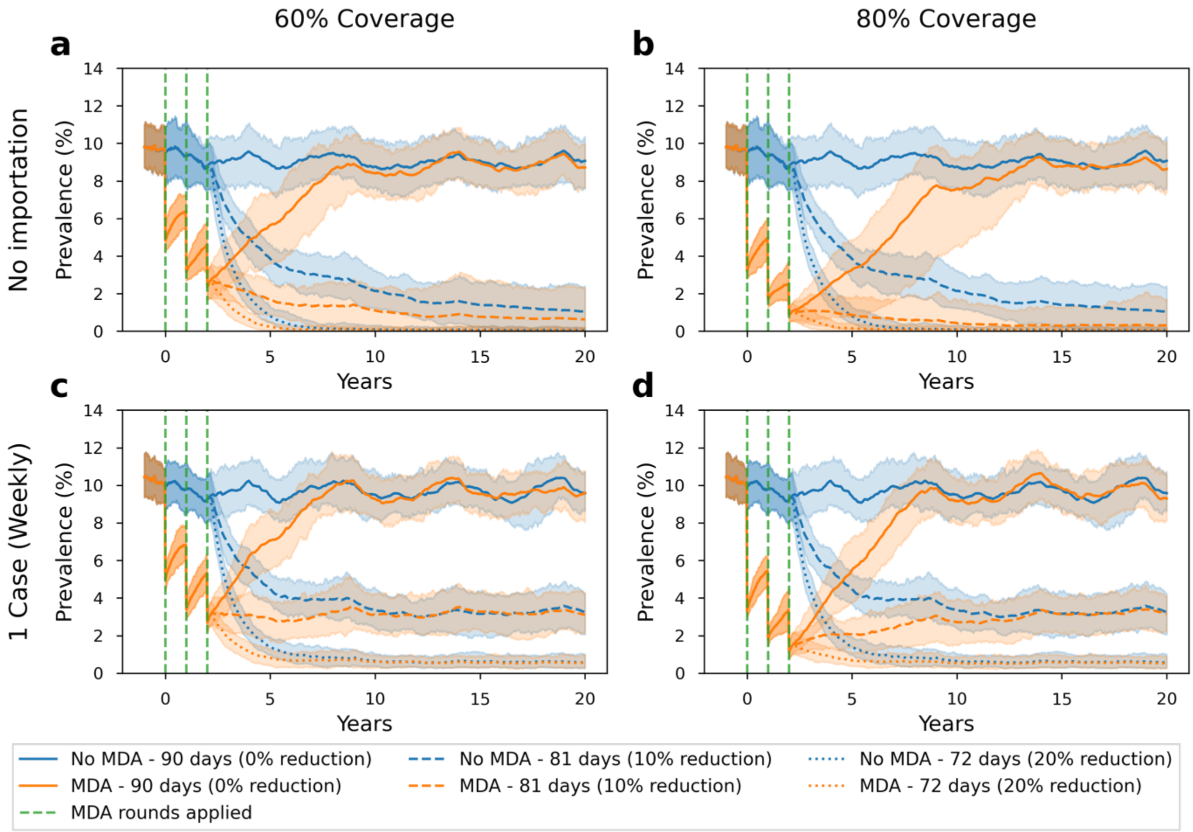

*Figure S17: The scabies prevalence in 20 years with (orange) and without (blue) MDA strategy in scenarios (a) 60% population coverage – no importation, (b) 80% population coverage – no importation, (c) 60% population coverage – with scabies importation, and (d) 80% population coverage – with scabies importation* ***with an average infestation duration of 90 days****. The green lines show when MDA rounds are applied (years 0, 1, and 2). Solid lines show scenarios with an average of 90 days (baseline – 0% reduction) duration of infestation throughout the simulation while dashed and dotted lines show an average of 81 (10% reduction) and 72 (20% reduction) days duration of infestation after year 2, respectively. MDA strategies are applied with random individual selection in 3 annual rounds. In scenarios with scabies importation, there is only one weekly scabies importation. Panel b represents results from Figure 3 in the main manuscript.*

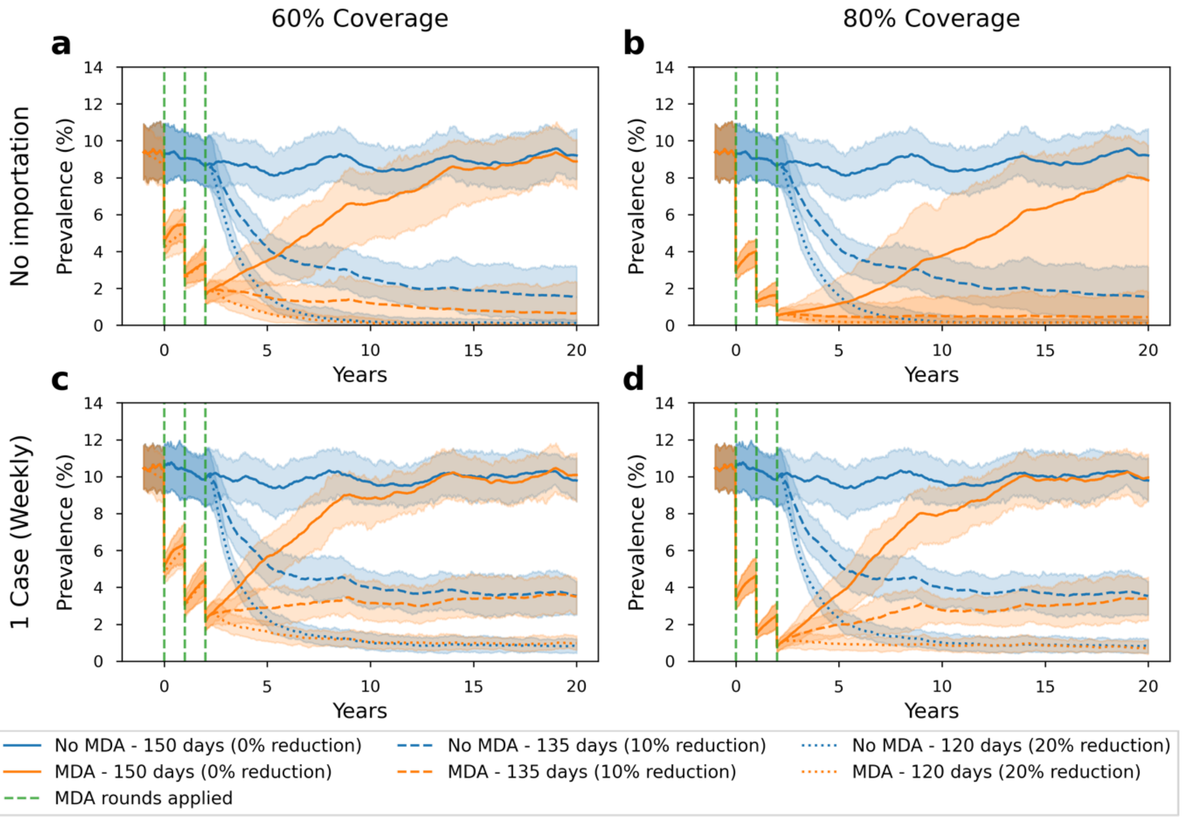

*Figure S18: The scabies prevalence in 20 years with (orange) and without (blue) MDA strategy in scenarios (a) 60% population coverage – no importation, (b) 80% population coverage – no importation, (c) 60% population coverage – with scabies importation, and (d) 80% population coverage – with scabies importation* ***with an average infestation duration of 150 days****. The green lines show when MDA rounds are applied (years 0, 1, and 2). Solid lines show scenarios with an average of 150 days (baseline) of the duration of infestation throughout the simulation while dashed and dotted lines show an average of 135 (10% reduction) and 120 (20% reduction) days of the duration of infestation after year 2, respectively MDA strategies are applied with random individual selection in 3 annual rounds. In scenarios with scabies importation, there is only one weekly scabies importation.*
